# Bias-free Regression Analysis in Nutrition (BRAiN) Index: A Predictive Model of Cognitive Function

**DOI:** 10.1101/2025.11.14.25340280

**Authors:** Federica Conti, Kevin Gluck, Andrea Stocco, Thomas Wood

**Affiliations:** Human Performance Center, Parker University, Dallas, TX 75229, USA; Institute for Human and Machine Cognition, Pensacola, FL 32502, USA; Department of Psychology, University of Washington, Seattle, WA 98195, USA; Institute for Learning and Brain Sciences, University of Washington, Seattle, WA 98195, USA; Department of Pediatrics, University of Washington, Seattle, WA 98195, USA

**Keywords:** Age-related cognitive decline, Dementia, Prediction model, Population data, Nutritional interventions

## Abstract

Evidence suggests the development of age-related dementia is strongly associated with numerous modifiable risk factors, particularly lifestyle behaviours such as diet, sleep, and physical activity. Yet, when exploring the dietary predictors of cognitive function, many scoring systems apply linear scales to eating patterns in a way that prioritizes simplicity over predictive accuracy. The present study employed a cross-validated elastic net regression algorithm on a large dataset of self-reported dietary data (n=28,968 individuals, mean age 56.3, age range 16-100) to identify the greatest nutritional predictors of participants’ performance on a validated online cognitive function test. Bias-free Regression Analysis in Nutrition (BRAiN) scores were computed, and their predictive power was found to outperform a well-recognized cognition-focused scoring system (Mediterranean-DASH Intervention for Neurodegenerative Delay, MIND) in held-out data. Non-linear relationships between intake level and cognition were noted for most foods, with greater consumption of animal and vegetable proteins, vegetables, nuts and seeds, and wholegrains, being positively associated with cognitive function, while the regular consumption of both refined fats and refined carbohydrates was negatively associated with cognitive function. Shifts in dietary patterns by age suggested that some relationships between diet and cognitive function in nutritional epidemiology may be confounded by age-related dietary trends. Our data-driven model can be easily adjusted over time to reflect potential shifts in dietary practices at the population level and inform relevant health authorities in the timely design of effective nutritional recommendations and guidelines to prevent or slow down the onset of age-related cognitive decline.

## Introduction

Age-related dementia is one of the major global health challenges of the 21^st^ century^1^. Dementia affects more than 57 million people worldwide,^2^ and its prevalence has steadily increased globally over the past 30 years^3^. Cognitive decline and dementia have non-modifiable risk factors such as age, sex, and genetic predisposition, yet recent enquiries strongly suggest that a large proportion of dementia cases may be preventable^4^. Epidemiological studies have revealed that certain lifestyle habits and behavioural practices, including diet, sleep, physical activity patterns, stress management, and the social and emotional support provided by meaningful interpersonal relationships, also play a prominent role in driving cognitive health outcomes^5–8^. Accordingly, the multi-dimensional aetiology of cognitive decline has called for a paradigm shift in the way we think of this chronic illness.

Statistical analyses on behavioural strategies to preserve cognitive health estimate that 45-72.6% of all dementia cases could be prevented through appropriate and timely interventions, including modification of lifestyle and improvements within the surrounding socio-economic environment^1, 9^. Where nutrition is concerned, the relationship between diet, nutrient status and dementia risk prompted the formulation of a potentially neuroprotective diet, known as the MIND (Mediterranean-DASH Intervention for Neurodegenerative Delay) diet^10^. The MIND diet is based on the results of the Memory and Aging Project, a US-based longitudinal study that took place between 2004 and 2013, involving 960 individuals. Participants were followed for an average period of 4.7 years via annual neurological assessments and food consumption questionnaires. Within the dietary surveys, 15 questions were selected to probe participants’ frequency of consumption of specific foods and food groups believed to be positively associated (e.g. berries, nuts, green leafy vegetables, olive oil) or negatively associated (e.g. red meats, butter, sweets, fast and fried foods) with cognitive health. A score of 0, 0.5 or 1 was assigned to each question and a total score, ranging from 0 to 15, was generated from the cumulative sum of these 15 individual components, with higher values describing dietary habits more likely to result in better cognitive outcomes^10^.

Since its inception, this MIND index has been extensively validated, and slower rates of cognitive decline have been observed in individuals following a MIND-aligned diet^11–17^. Nonetheless, some conflicting or inconclusive results have also emerged, especially in the context of certain comorbidities such as being overweight or obese. For example, Barnes et al. (2023) conducted a three-year randomized controlled trial in 604 cognitively unimpaired older adults with a body mass index greater than 25 and a family history of dementia. Half of the participants were assigned to the MIND diet, while the other half were assigned to a control diet involving mild caloric restriction. Both groups showed improvements in global cognition from baseline to year 3, while brain MRI outcomes, including white-matter hyperintensities, hippocampal volumes, and total grey-and white-matter volumes were not significantly different between the intervention and control arms^17^.

One of the primary issues with the real-life applicability of the MIND score and similar dietary scoring systems is the mathematical relationship between the consumption of certain foods and cognitive metrics. In this framework, component scores are established based on numerical scales that have been somewhat arbitrarily defined, where different frequencies of consumption are often assumed to be equidistant to one another while also affecting cognitive function in a linear fashion. As an example, in the MIND framework, consuming “less than 1 serving of cheese per week”, “between 1 and 6 servings per week”, and “more than 7 servings per week”, are assigned a numerical score of 1, 0.5 and 0, respectively, thus implying that the cognitive benefit of consuming less than 1 serving per week is cut in half when consuming anywhere between 1 and 6 servings, and completely disappears when exceeding 6 servings per week. However, whether empirical evidence supports these mathematical assumptions is unclear.

A novel methodological approach to counteract these issues would be to identify key nutritional variables based on a larger, ideally more geographically and culturally diverse dataset, without assuming any pre-existing relationship between dietary intakes and cognitive outcomes. To accomplish this goal, we developed an elastic net regression model to elucidate the relative contribution of common foods and food groups to cognitive function and thereby define a data-driven nutrition score. We built and validated our model using the Food for the Brain (FFB) database and compared the resulting nutrition score to an approximated MIND score generated from the same database, based on the queries at our disposal. We describe the theoretical formulation and implementation of our statistical model and discuss its performance, relevance, and applicability in the context of dementia prevention.

## Methods

### Data collection

Data used for the present study were collected between November 2022 and June 2025 through the online Cognitive Function Test (CFT) developed by the Food for the Brain Foundation (https://foodforthebrain.org/). This short, self-administered, computerized test was designed to provide a quick and reliable assessment of cognitive abilities and has been validated against standard paper-based tests of episodic memory, processing speed, and executive function^18^. A small pilot trial has further confirmed the internal consistency and reliability of the online version of this screening tool for large-scale use^18^. Following the cognitive function test, respondents completed an optional comprehensive health and lifestyle questionnaire comprised of 144 questions about their medical history, sleep, dietary and physical activity behaviours, as well as social, psychological and emotional well-being. Our current analysis focuses on the 21 questions that make up the dietary subset of the questionnaire and the extent to which respondents’ answers to those questions predict their performance on the cognitive function test.

A total of 31,266 eligible participants voluntarily took the online cognitive function test in the study time window. The dataset obtained from this cohort included respondents’ basic demographic information (age, gender, country of origin, and years of education), their cognitive test score, and their answers to the health and lifestyle questionnaire.

### Data management and human research ethics

Sensitive data collected during the cognitive function test included only participants’ first name and age. Upon completion of the cognitive function test and the health and lifestyle questionnaire, participants were assigned a 32-digit alpha-numeric identifier, and their score, answers and demographic details were stored exclusively against this number. De-identified data from the Food for the Brain database were then extracted and transferred to a REDCap database (Vanderbilt University). Based on these data management procedures, subjects could not be identified from their responses. Therefore, the University of Washington Institutional Review Board confirmed that this work did not constitute human subjects research and ethical approval was not necessary for the conduct of the present study.

### Data cleaning and preprocessing

Prior to formal analyses, data were carefully screened for missing demographic data, excessive missing entries (e.g. participants completing only the demographic portion of the survey or skipping more than 50% of the survey questions), formal errors (e.g. years of education exceeding age) and inconsistencies (e.g. between primary and follow-up questions). These instances were found to occur in a relatively small group of responses (4.79%) and all data provided from the participants in question were excluded from the dataset. Next, we selected respondents who were at least 16 years of age and had a cognitive function score of at least 15 out of 100, the minimum that is realistically possible from somebody meaningfully engaging with the test. Reported years of education were truncated at the upper end of the education range based on the assumption that respondents would likely not have completed more than 25 years of full-time education. Finally, isolated missing entries in one or more of the nutrition-related questions were identified in 19 respondents. These instances were also removed. A flow diagram outlining the steps of this data cleaning process can be found in Supplementary Figure 1.

### Dietary queries and restructuring

Dietary queries from the FFB health and lifestyle questionnaire probed participants’ consumption of specific foods and food groups, including fruits, vegetables, wholegrains, animal and vegetable proteins, tea, coffee, wine, table sugar and sweets, fried and fast foods, refined carbohydrates and sunflower oil (the primary vegetable oil used in the United Kingdom). Participants estimated their frequency of consumption against 5 possible answers, increasingly ordered (e.g. “Never/Rarely”, “1-2 a week”, “Every other day”, “Once a day”, “2 or more a day”). Additionally, participants were also asked to indicate whether they were following a vegan diet (“Yes”, or “No”). The complete list of original nutrition questions is presented in Supplementary Table 1. Preliminary analyses of the distributions of participants’ responses revealed that several response options were selected by a relatively small proportion of participants (i.e. <10%, see Supplementary Figure 2). To avoid potential biases in subsequent statistical analyses caused by the presence of heavily skewed distributions, each question’s answers were regrouped to reduce question-level categories from five to three (see Supplementary Table 2). Additionally, given that three pairs of questions probed participants’ consumption of very similar food items (i.e. refined carbohydrates and bread/biscuits/pasta/pizza, fish and oily fish, dark leafy and cruciferous vegetables), these pairs were combined into single questions to eliminate collinearity.

### Statistical model and analyses

Statistical analyses were performed using R Studio (R Core Team, 2024, version 4.4.2). Prior to data analysis we split our dataset into two subsets for training (70%) and testing purposes (30%), respectively. We built an elastic net regression (ENR) model using the R package *glmnet*^19^ to predict cognitive function scores using age, gender, years of full-time education and the 21 nutrition-related questions of the health and lifestyle questionnaire as predictors.

Elastic net regression combines two linear regression techniques, LASSO and Ridge, to handle multicollinearity issues between model predictors, selecting the appropriate regression coefficients that best apply to the dataset under investigation, and reducing both the occurrence of prediction errors and the probability of overfitting^20^. The algorithm is designed to regularize the mathematical model by shrinking redundant or irrelevant regression coefficients towards zero, thus selecting only predictor variables found to exert significant effects on the outcome measure. To justify this methodological choice, we first validated our model against a classical linear regression algorithm and conducted a sensitivity analysis of the alpha level between 0 and 1 by 0.1 increments. Results from this direct comparison pointed to a strong agreement between the methods, as indicated by very similar regression coefficients across the whole spectrum of selected variables and regardless of the alpha level, with overall marginally smaller mean-squared errors and larger R-squared statistics favouring the elastic net regression. A breakdown of this comparative analysis is presented in Supplementary Tables 3 and 4. We then set the alpha level at 0.5 and imposed a 10-fold cross validation strategy on the training phase of our model development. The shrinking parameter we used to fit our model (i.e. lambda) was selected by minimizing the mean squared error (MSE) of the predicted values in the 10-fold cross-validation procedure. Since questionnaire responses were not assumed to be in linear relationships with the outcome metric, categorical predictors were converted into individual dummy variables for each level of response. This approach naturally increased the number of predictors, yielding a total of 66 variables. Lastly, on each application of the ENR mentioned throughout this work, model performances were evaluated by computing and comparing the MSE obtained on the validation dataset to the baseline MSE, and we used the R^2^ statistic (Olkin-Pratt estimator), or coefficient of determination, to assess how well each model was able to explain the variance observed in our outcome variable (CFT score). Following common practice, for all statistical analyses performed, the threshold for statistical significance was set at 0.05.

### MIND* Scores calculation

The first question we address is whether a data-driven, ENR-based approach to computing MIND scores outperforms the traditional fixed linear MIND scores. Therefore, we constructed a modified MIND score, referred to as a *MIND* Linear* score, by selecting the survey questions that most closely resembled the 15 queries featured in the original MIND diet model and by scoring the corresponding answers using analogous linear scales. Given that it was not possible to find an equivalent question that would perfectly match each original question, our MIND* Linear scores were calculated from 10 questions with final scores ranging from 0 to 9.5 (see Supplementary Table 5). We then used the same 10 variables as predictors in our ENR model and computed individual data-driven nutrition scores, referred to as *MIND* ENR* scores. These scores were derived from the weighted sum of participants’ answers to the selected questions, i.e. the dot product of the dummy variables corresponding to the answers they had selected and their corresponding regression coefficients. For ease of comparison and analytical consistency, we then scaled the resulting scores to the range 0-9.5. To compare the predictive power of these two dietary indices, we ran separate Spearman’s correlation analyses with participants’ CFT scores. Additionally, we extracted the regression coefficients and relative weights obtained from the ENR to characterise the mathematical relationships within variables suggested by the data-driven algorithm against the linear scales employed in the original MIND diet model. MSE and R^2^ statistics were computed to evaluate our model’s overall performance.

Lastly, we examined the relationship between significant dietary predictors identified by the ENR algorithm and cognitive function in the context provided by prior evidence on the neuroprotective or harmful effects of specific foods and food groups. When counterintuitive findings emerged, we employed propensity-score methods to reduce potential confounding by demographic and lifestyle factors. In these scenarios, separate logistic regression models were fitted to estimate each participant’s probability of being a high consumer of the food or food group in question (e.g. fish, ≥3 times per week) conditional on age, sex, education, and relevant supplement use (e.g. omega-3), if applicable. Inverse probability of treatment weights derived from these models were used in subsequent weighted linear regressions of cognitive function on dietary intake, providing adjusted mean differences in cognition associated with high dietary intake.

### BRAiN Scores calculation

Next, we ran a more comprehensive ENR by including all 21 nutrition-related queries in the FFB dataset, as well as additional demographic predictors such as age, gender, and years of full-time education. The regression coefficients were then used to calculate the relative contribution of each demographic and nutrition question (as well as the contribution of each possible answer within each question) to the cognitive function score. Participants’ extended nutrition scores, referred to from this point forward as BRAiN (Bias-free Regression Analysis in Nutrition) scores, were computed using the same mathematical procedure previously described. Here again, we ran Spearman’s correlation analyses between participants’ BRAiN scores and cognitive function scores. By comparing the resulting correlation coefficient to the one obtained from the previous, MIND-based approach, we were able to assess whether the inclusion of a greater number of nutrition-related variables, alongside demographic data, would improve the predictive power of our regression model. MSE and R^2^ statistics were computed to evaluate the performance of the BRAiN model.

Subsequently, we conducted a series of exploratory analyses to understand the impact of key demographic variables (i.e. age and gender) on both cognitive function and nutrition scores. Where age is concerned, we conducted a qualitative analysis to investigate potential non-linear relationships with both BRAiN and cognitive scores. A decline in both scores was expected to occur with increasing age^21, 22^, so we generated regression lines for both scores using Locally Estimated Scatterplot Smoothing (LOESS) and employed segmented regression analyses to estimate breakpoints along each curve, denoting sharp and meaningful downward deflections. We then used non-parametric bootstrapping and permutation tests to directly compare the slopes of the regression lines past their respective breakpoints.

### Regression analyses based on age and age groups

To evaluate whether the relative contributions of nutrition-related predictors to respondents’ cognitive function scores would differ based on participants’ age, we built an additional ENR model by introducing interaction terms between age and nutrition-related predictors. We explored the statistical significance of these interactions and looked at potential differences in the most significant predictors compared to our previous model to gain a better understanding of how certain dietary trends may shift as age increases, and how such changes may in turn be positively or negatively associated with changes in cognitive function.

Lastly, to evaluate whether the BRAiN index allows for more accurate predictions based on age group, we stratified our training and testing datasets into three participant groups based on the breakpoints identified by the segmented regression algorithm described above (16–43-year-olds, 44-68-year-olds and older-than-68, cf. section *BRAiN scores*) and computed the MSE and R^2^ statistics obtained from each age-specific regression model. Differences in top predictors of cognitive functions within each dataset were also investigated.

## Results

### Participants’ characteristics

Table 1 provides a summary of participants’ basic demographic characteristics and corresponding descriptive statistics (means and standard deviations or counts and percentages, as appropriate). A total of 28,968 participants were retained in the final dataset for this study. Of these, approximately two thirds (65.3%) were female and one third were male (34.7%). Half of the participant sample (51.7%) was between 51 and 70 years and mean age was 56.3. Years of full-time education ranged from 0 to 25, with an average value of 14.5 years. A subset of 23,238 participants further provided information regarding their country of origin. Within this group, most participants came from the United Kingdom (47.93%), The United States of America (21.35%), Australia (6.54%) and Canada (6.35%). The remaining 17.83% of this subsample came from a cumulative pool of 135 countries, with 79 of these countries including fewer than 10 participants each (see Supplementary Figure 5 for a world map of study participants who provided cultural background data).

**Table 1.**
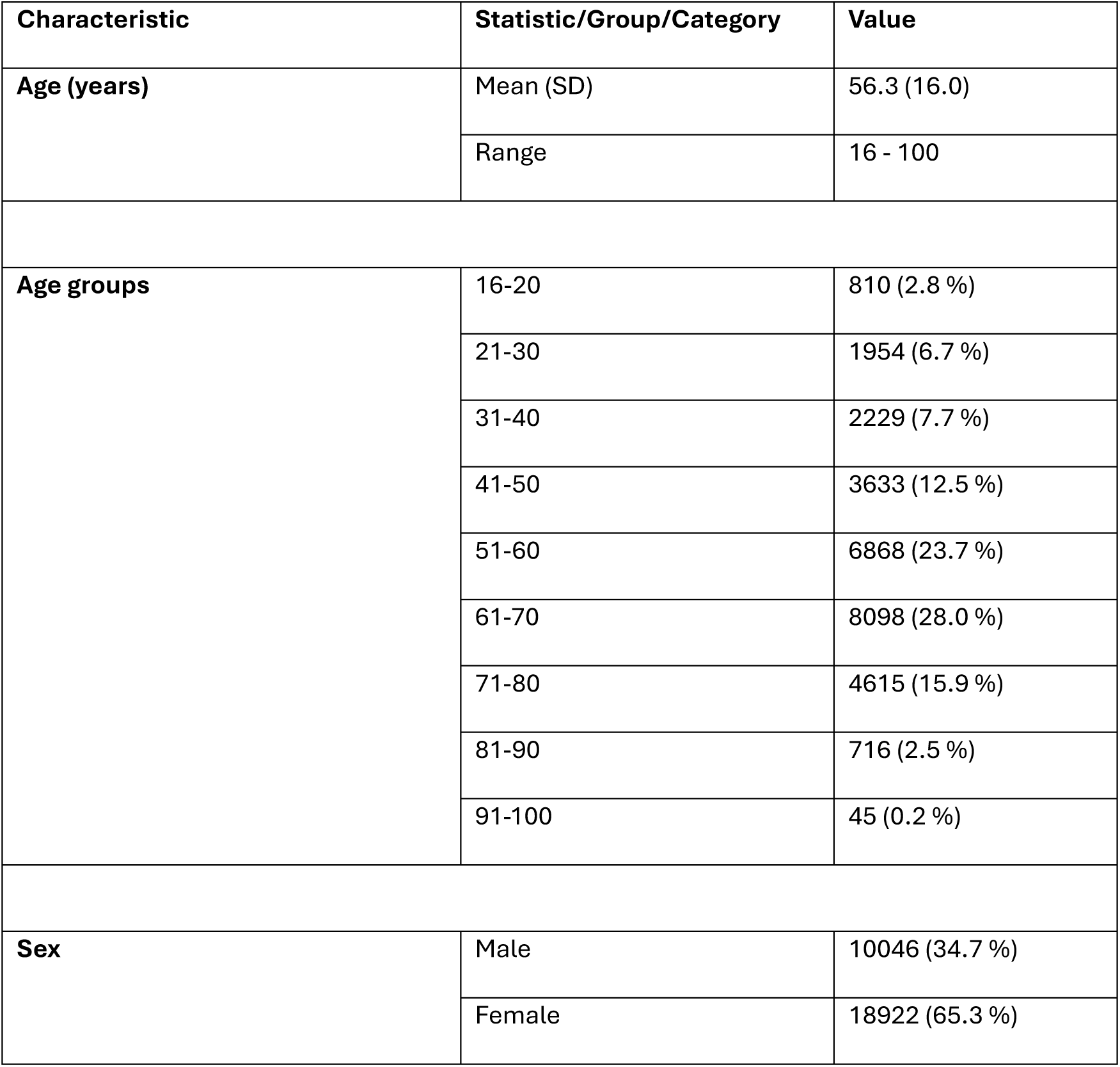
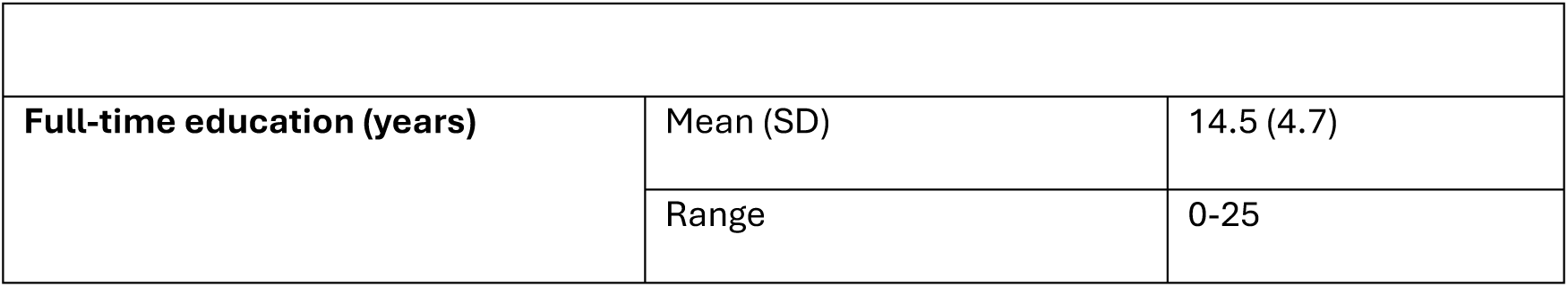
Participants’ demographic characteristics and corresponding descriptive statistics.

### Evaluation of MIND* scores

We begin by comparing MIND* scores calculated under the linear assumptions of the original MIND model (MIND* Linear, questions on a 0-1 scale) to scores derived from the ENR algorithm (MIND* ENR) to determine whether the data-driven, ENR-based approach allows for better predictive accuracy in terms of participants’ cognitive function. Using the same responses to the MIND questions, the ENR rmodel reduced the MSE of the CFT score prediction in the validation dataset from 156.35 to 154.46 (R^2^ = 0.01). Descriptive statistics for both participants’ MIND* Linear and MIND* ENR scores are summarised in Table 2, while their relationship with participants’ CFT scores is illustrated in Figure 1 using density plots. Smoothed trend lines were estimated using locally estimated scatterplot smoothing (LOESS) to capture general patterns without assuming linearity. Shaded areas represent 95% confidence ribbons.

**Figure 1.**
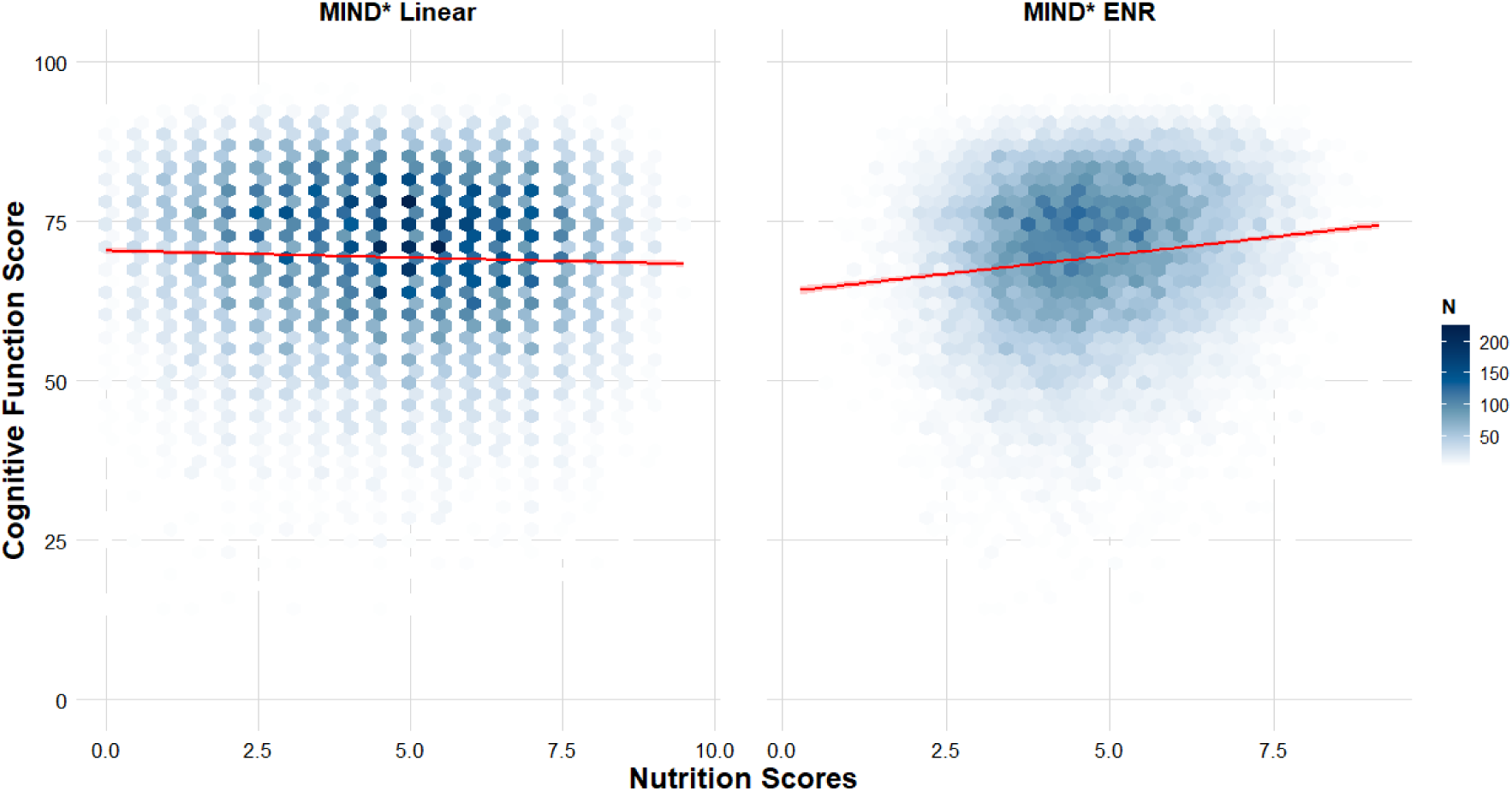
Density plots and LOESS-smoothed relationships between participants’ cognitive scores from the online Cognitive Function Test and both MIND* Linear and MIND* ENR nutrition scores. MIND* Linear scores were obtained by selecting the survey questions that most closely resembled the 15 queries featured in the original MIND diet model and by scoring the corresponding answers using analogous linear scales (see Supplementary Table 5). Scores were calculated from 10 dietary queries and ranged from 0 to 9.5. The same 10 queries were used as predictors in the elastic net regression model and MIND* ENR scores were derived from the weighted sum of participants’ answers, i.e. the dot product of the dummy variables corresponding to the answers participants had selected and their corresponding regression coefficients. For ease of comparison and analytical consistency, MIND* ENR scores were rescaled to the range 0-9.5.

**Table 2.** Descriptive statistics for participants’ MIND* scores and data-driven scores.

|  | **Minimum** | **1st quartile** | **Median** | **Mean** | **SD** | **3rd quartile** | **Maximum** |
| --- | --- | --- | --- | --- | --- | --- | --- |
| **MIND\* Linear** | 0 | 3.00 | 5.00 | 4.69 | 2.02 | 6.00 | 9.50 |
| **MIND\* ENR** | 0.28 | 3.75 | 4.64 | 4.73 | 1.37 | 5.70 | 9.12 |

Spearman’s correlation analyses revealed a small, negative association between participants’ MIND* Linear scores and CFT scores (correlation coefficient: *r* =-0.05, *p* < 0.001). By contrast, a positive association was found between participants’ MIND* ENR scores and CFT scores (correlation coefficient: *r* = 0.13, *p* < 0.001), suggesting better ecological validity of the ENR-based approach.

Although increasing frequency of consumption generally reflected greater impact on participants’ cognition, regression coefficients in the MIND* ENR model were not linearly related, as they were not equidistant to each other (see Figure 2). In other words, if specific food intakes were found to be positively or negatively associated with CFT, additional intake was not associated with a linear change in predicted CFT score of a similar magnitude.

**Figure 2.**
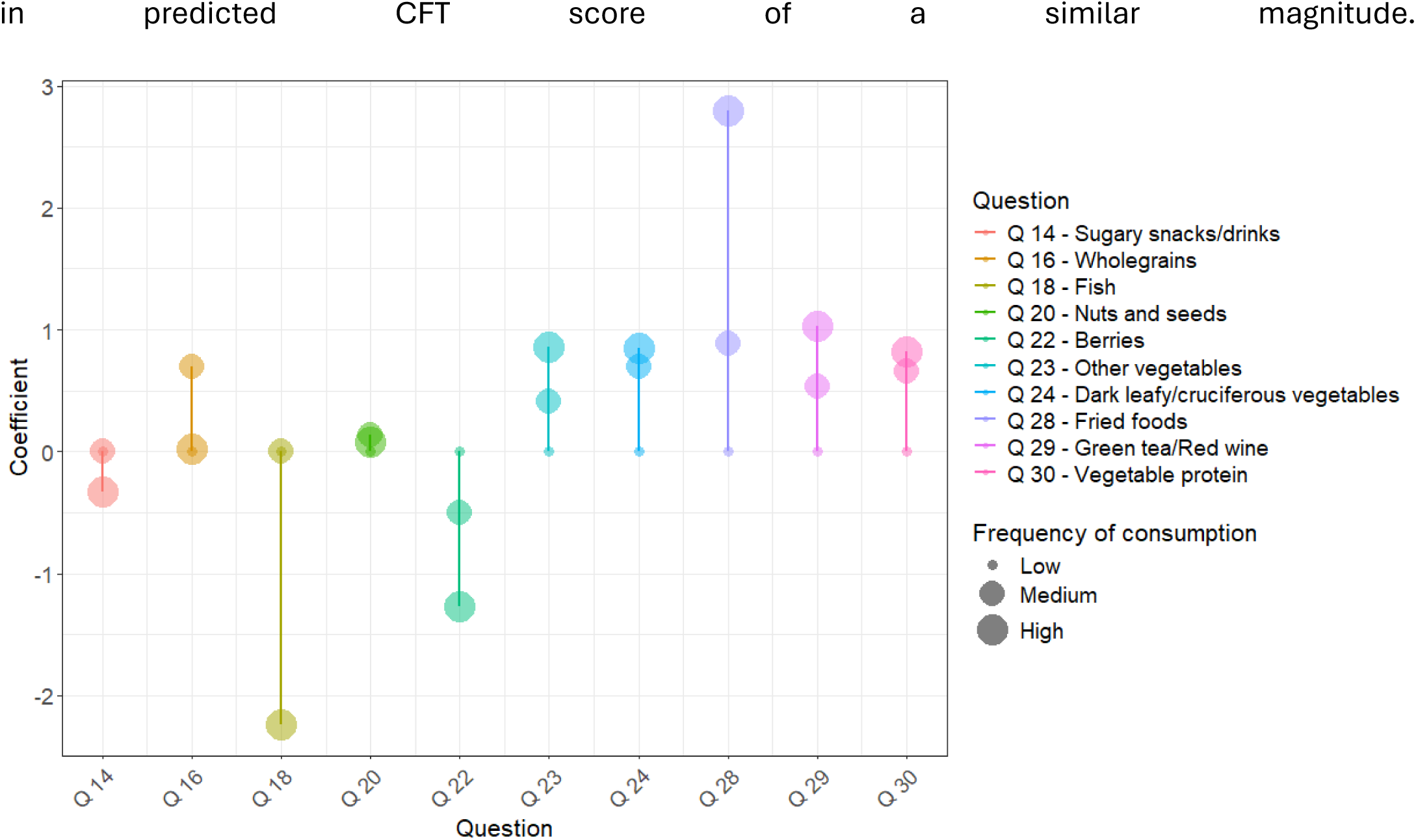
ENR regression coefficients for significant predictors of cognitive function using the nutrition questions selected by the MIND model. Significant predictors are grouped by question. Regression coefficients are displayed as full dots and joined by straight lines to show unequal distances between possible answers within the same nutrition-related question. Within each question, dot sizes, from smaller to larger, reflect increasing frequency of consumption (e.g. “None/Rarely”, “1 a day or less”, “2+ a day”) to showcase that the relationship between dietary intake and cognition usually is not linear. Question numbers reflect the number from the original FFB health and lifestyle questionnaire.

The complete list of ENR regression coefficients, grouped by question, is presented in Table 3. These estimated contributions indicate that regular consumption of wholegrains, vegetables (of all kinds), vegetable protein and, to a smaller extent, nuts and seeds, was positively associated with cognitive function. Green tea and red wine were also positively associated with CFT score, though as they were combined into the same question in the FFB survey, the extent to which this effect may be driven by one or the other, or a combination of both, is unclear. By contrast, both sugary snacks or drinks and fish intake were negatively associated with CFT score, although this last observation may have resulted from the relatively low percentage of responses allocated to the higher category of fish intake (Supplementary Figure 3). Lastly, berries and fried foods were found to be negatively and positively associated with CFT score, respectively. Here, socio-economic factors such as income, food affordability and convenience, may have come into play, as suggested by the distribution of participants’ responses to these selected questions by age group (i.e. 16-40, 40-70, 70+ year olds, Figures 3-5) and education group (see Supplementary Figures 6-8).

**Figure 3.**
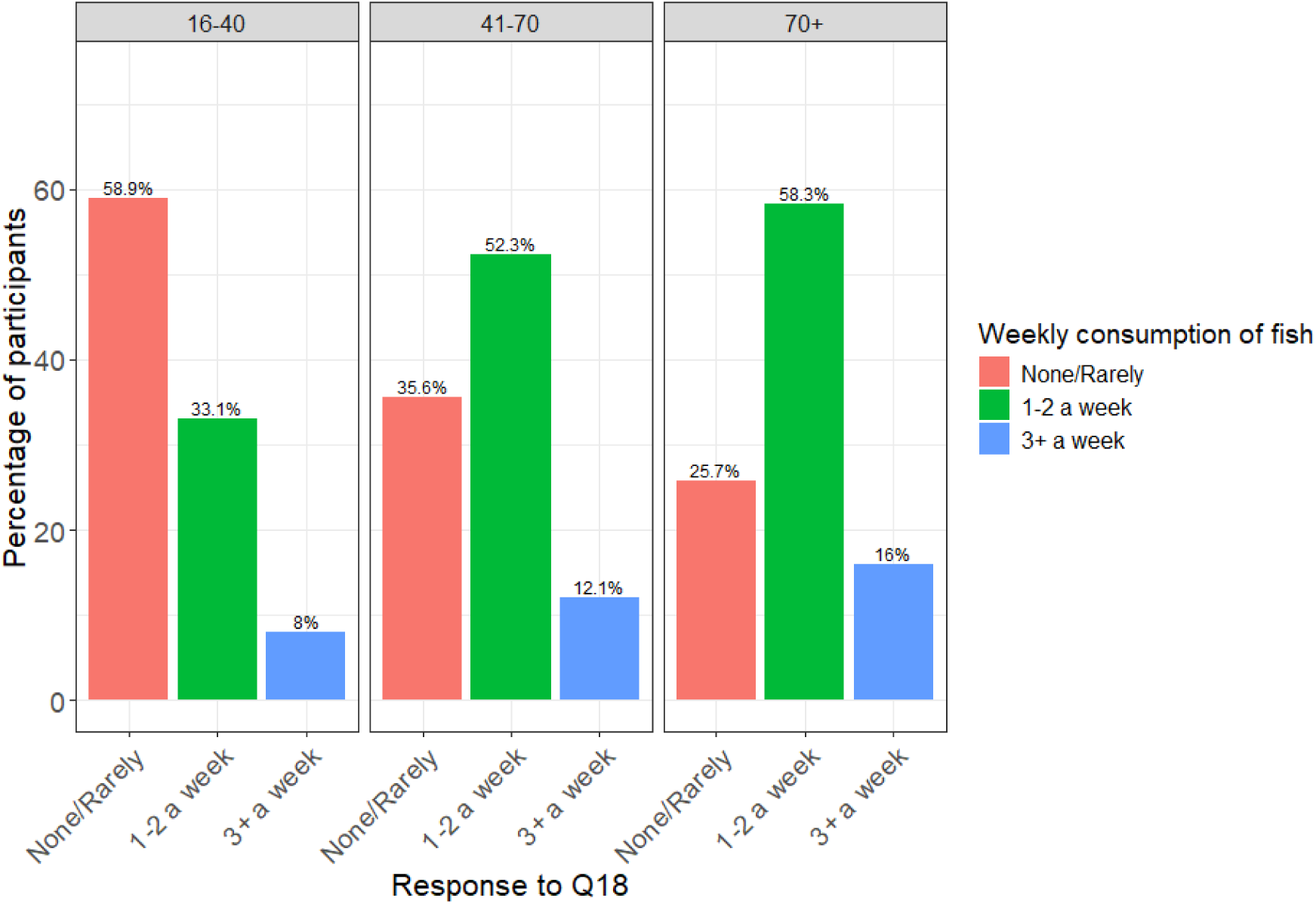
Distribution of participants’ responses to questions 18 (fish) by age group.

**Figure 4.**
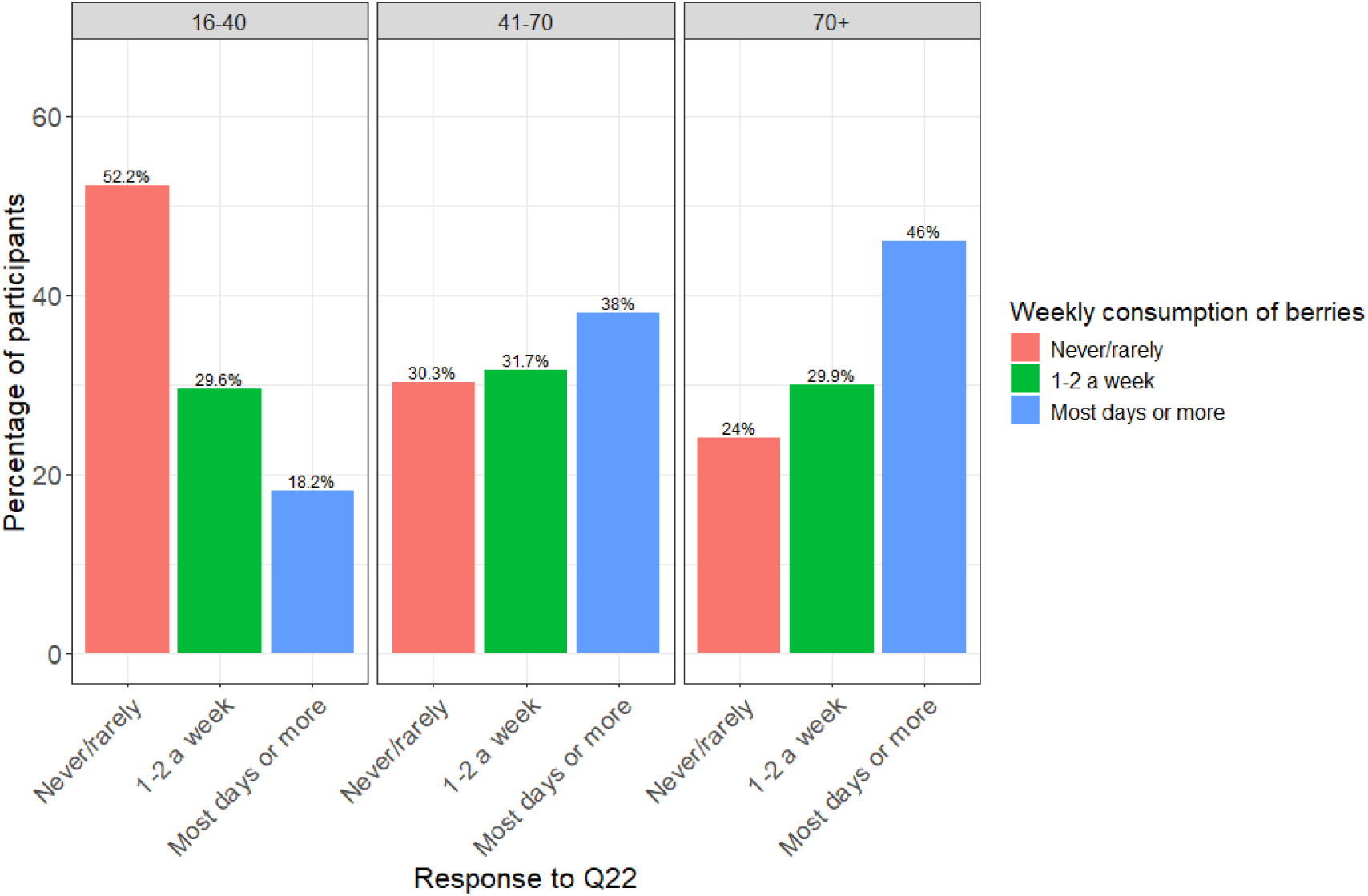
Distribution of participants’ responses to questions 22 (berries) by age group.

**Figure 6.**
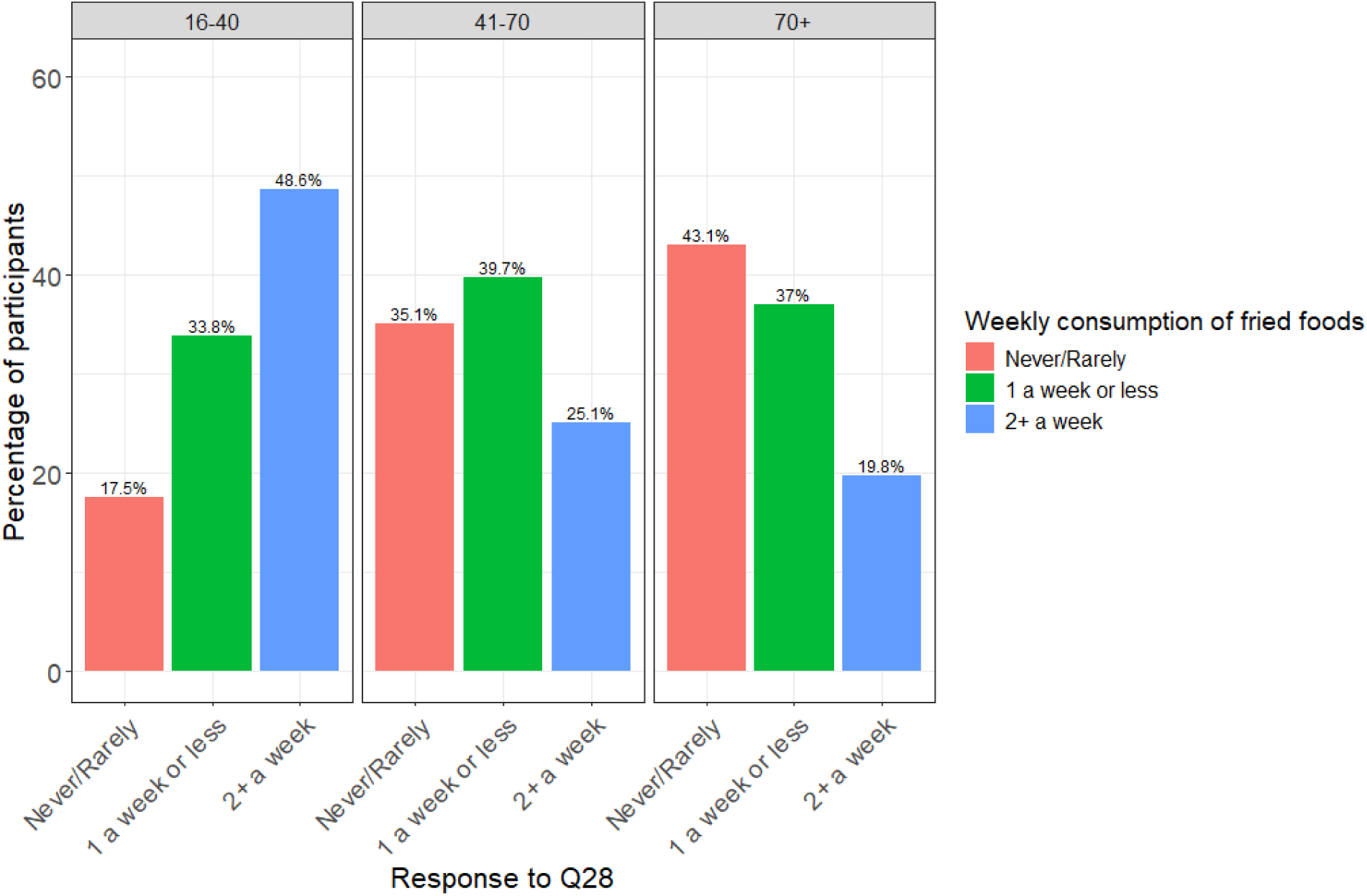
Distribution of participants’ responses to questions 28 (fried foods) by age group.

**Table 3.** ENR regression coefficients for predictors of cognitive function grouped by question.

| Question | Possible answers | Regression coefficients |
| --- | --- | --- |
| **Q14:** How many sugar-based snacks or drinks (biscuits, cakes, chocolate, sweets, fizzy drinks, fruit juices) do you consume each day? | None/Rarely<br>1 a day or less<br>2+ a day | 0<br>0<br>-0.33 |
| **Q16:** How many times a week do you eat wholegrains (e.g. brown rice, oats, wholegrain bread, wholewheat pasta)? | 1 a week or less<br>2-3 a week<br>More than 3 a week | 0<br>0.70<br>0.02 |
| **Q18:** How many times a week do you eat fresh oily fish (e.g. salmon, mackerel, sardines, herring)? | None/Rarely<br>1 to 2 a week<br>3+ a week | 0<br>-1.33<br>-2.24 |
| **Q20:** How frequently do you eat raw walnuts, pecans, macadamia, chia or flax seeds? (a handful or small cup) | Never/Rarely/Occasionally<br>2-3 a week<br>More than 3 a week | 0<br>0.14<br>0.08 |
| **Q22:** How many servings of berries, cherries, plums, pomegranate or their juice do you have a day? | Never/Rarely<br>1-2 a week<br>Most days or more | 0<br>-0.51<br>-1.27 |
| **Q23:** How many servings of orange or red vegetables do you have a week (e.g. carrot, beetroot, sweet potato, squash, peppers)? | 1 a week or less<br>2-4 a week<br>5+ a week | 0<br>0.41<br>0.86 |
| **Q24:** How many times a week do you eat dark green or cruciferous vegetables? (e.g. broccoli, cabbage, cauliflower, brussels sprouts)? | 2 or less a week<br>3-4 a week<br>5 or more a week | 0<br>0.70<br>0.85 |
| **Q28:** How many times a week do you eat fried, deep fried or browned foods including crisps, chips and fried take away food? | Never/Rarely<br>1 a week or less<br>2+ a week | 0<br>0.88<br>2.79 |
| **Q29:** How many green or herbal teas or small glasses (175mls) of red wine do you drink a week? | None/Rarely<br>Regularly<br>7+ a week | 0<br>0.54<br>1.03 |
| **Q30:** How many times a day do you eat vegetable protein (e.g. beans, lentils, tofu, quinoa, peas, corn)? | None/Rarely<br>1 or 2 a week<br>More than 3 a week | 0<br>0.66<br>0.82 |

**Table 4.** Descriptive statistics for participants’ BRAiN scores.

| Minimum | 1st quartile | Median | Mean | SD | 3rd quartile | Maximum |
| --- | --- | --- | --- | --- | --- | --- |
| 0 | 51.86 | 63.62 | 61.74 | 15.68 | 73.47 | 100 |

Considering that both berries and fish are recognised as neuroprotective within the relevant literature due to their high content of antioxidants and omega-3 fatty acids, respectively, we investigated whether respondents consuming lower amounts of berries or fish were resorting to use of supplements instead (for which specific questions were also included in the health and lifestyle questionnaire). Spearman’s correlation analyses by age group (i.e. 16-40, 40-70, 70+ year olds) revealed small, yet statistically significant positive associations between the consumption of berries or fish on the one hand, and regular supplementation with antioxidants and vitamin C, or omega-3 fatty acids on the other (all 0.06 ≤ *rho* ≤ 0.15, all *p*-values <.001). Supplementation was also found to increase steadily with age, likely due to greater awareness of the increased risks associated with declining cognitive function, as well as greater willingness and financial capability to supplement with nutrients offering evidence-based cognitive and health benefits.

In the attempt to better understand the contribution of berries and fish consumption to cognitive function in the broader context of respondents’ demographic characteristics and chosen supplementation strategies, we performed propensity score analyses including both demographic variables (age, gender, years of education) and relevant supplement use (fish – omega-3 fatty acids; berries - vitamin C + antioxidant). For fish, results indicated that participants who consume fish 3 or more times per week had cognitive scores approximately 0.6 points lower than those who ate fish less frequently (95 % CI: [–0.9, –0.3], *p* = 0.0001), so the negative association between fish intake and CFT score held even after controlling for confounding factors.

By contrast, participants who consume berries most days of the week or more had cognitive scores approximately 0.6 points higher than those who ate them less frequently (95 % CI: [0.3, 0.86], *p* = 0.0001), thus indicating that, once potential confounding factors are accounted for, higher berry intake tends to be associated with higher CFT scores.

### The BRAiN Model

By selecting all nutrition-focused variables from the original FFB survey alongside key demographic variables (age, gender, and years of full-time education), our extended ENR model reduced the baseline MSE of predicted CFT score in the validation dataset from 156.35 to 132.17 (R^2^ = 0.15).

Figure 7 presents a graphical representation of the top 25 identified nutrition question predictors and their respective regression coefficients, in decreasing order of absolute value. The numerical values of these coefficients are displayed, with the direction and colour of each horizontal bar in the graph reflecting the predictor’s positive (right, green) or negative (left, red) contribution to cognitive function. Importantly, the regression coefficient attributed to age was multiplied by 10 to reflect the contribution of this demographic variable to cognitive function by decade rather than year.

**Figure 7.**
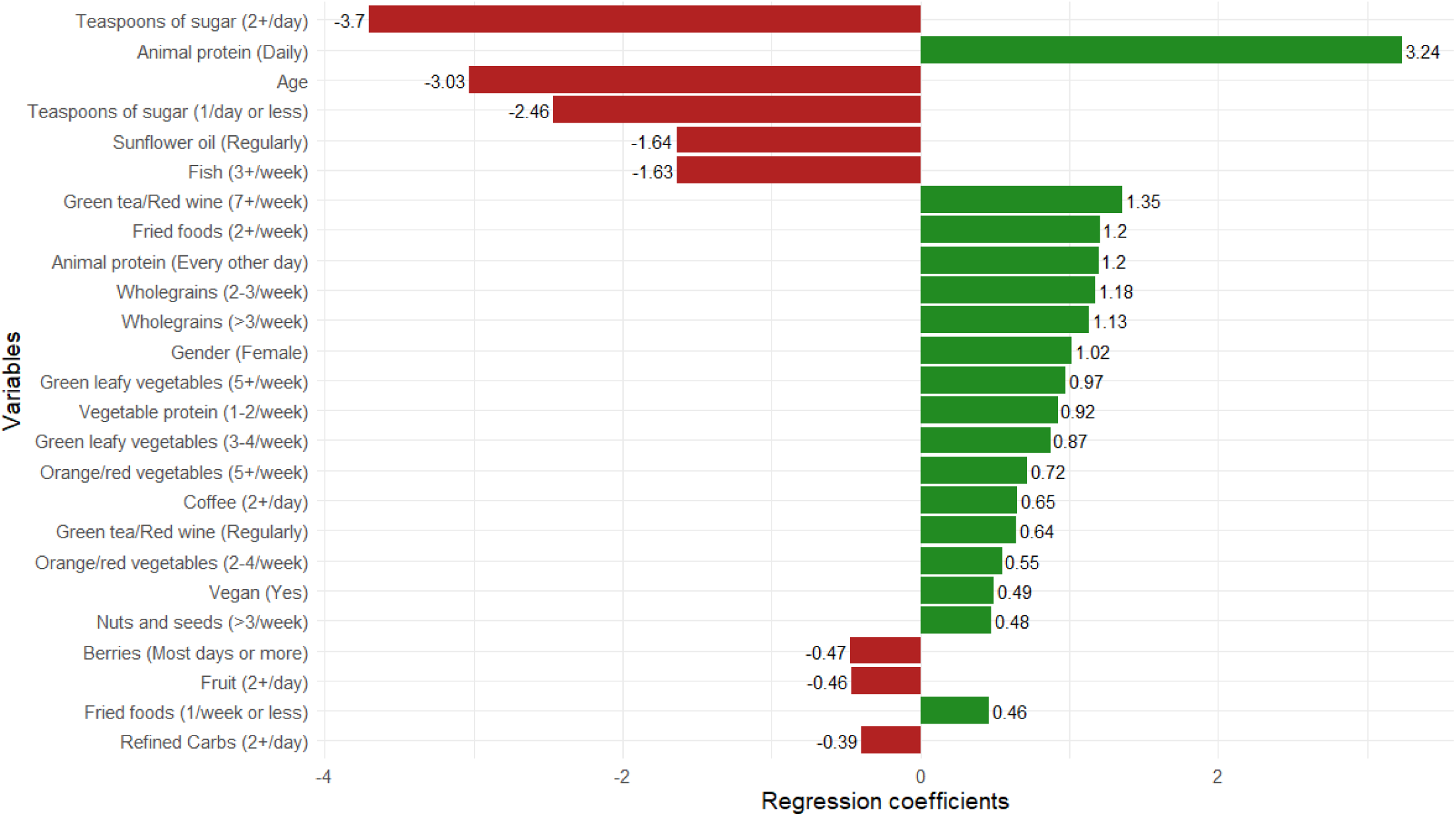
Top 25 predictors of cognitive function identified by the extended ENR algorithm, including all 21 dietary queries from the health and lifestyle questionnaire, as well as demographic variables (i.e. age, sex, and years of education). Identified predictors correspond to dummy variables, i.e. specific frequencies of consumption within each dietary question. The numerical values of each regression coefficient are displayed, with the direction and colour of each horizontal bar in the graph reflecting the predictor’s positive (right, green) or negative (left, red) contribution to cognitive function. The regression coefficient attributed to age was multiplied by 10 to reflect the contribution of this demographic variable to cognitive function by decade rather than year.

Next, we combined the dummy variables corresponding to the three possible answers to each question back to the original predictors to compute the cumulative contribution of each demographic or nutrition-related query to cognitive function. The relative weights obtained from this analysis are displayed in Figure 8.

**Figure 8.**
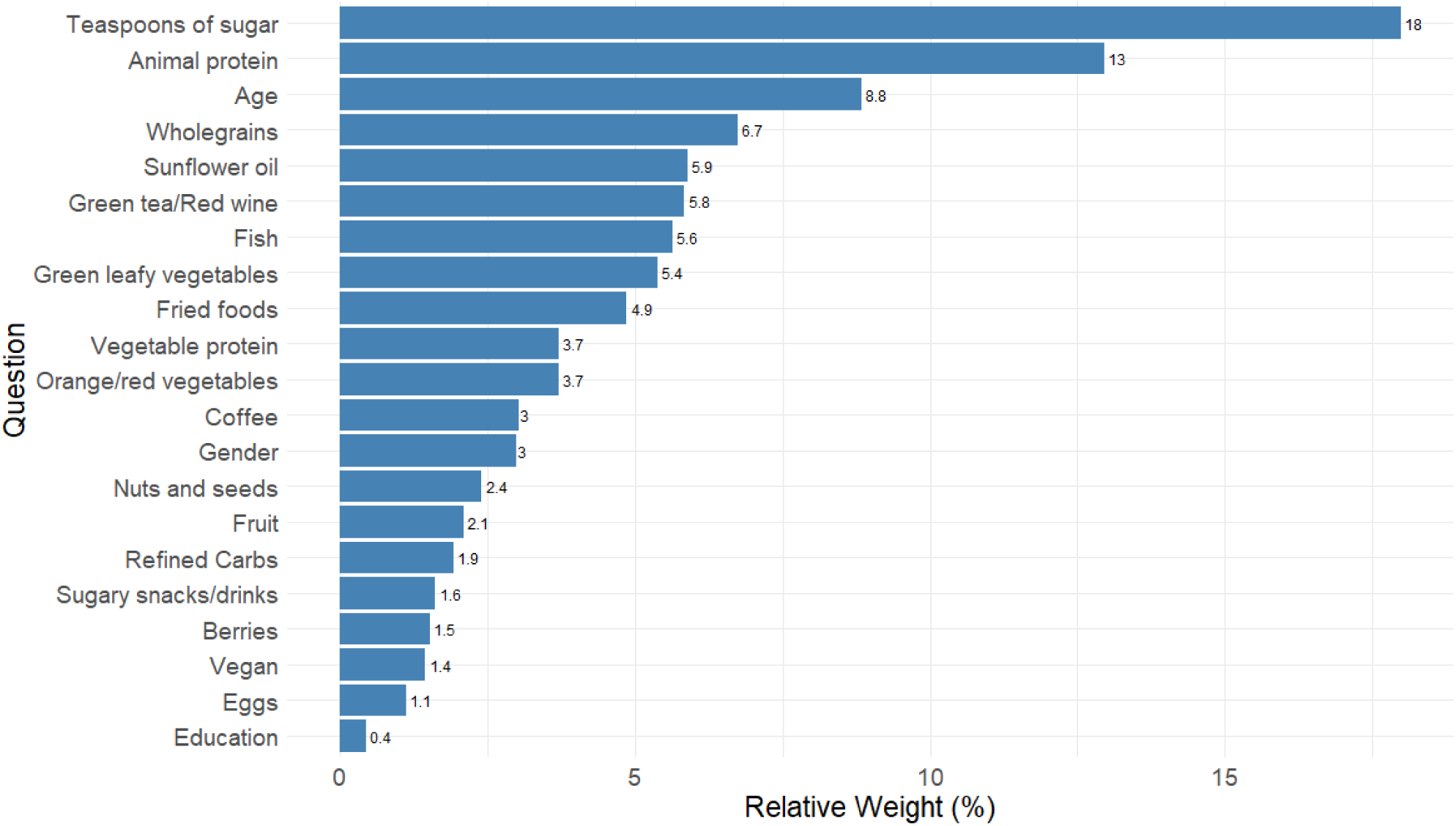
Relative contributions of demographic and nutrition-related questions to cognitive function. Relative contributions for each question are obtained from the sum of the regression coefficients attributed to the corresponding dummy variables, in absolute values. As such, these contributions reflect the overall impact each dietary query has on participants’ cognitive function scores, regardless of the positive or negative effects associated with specific frequencies of consumption within each food group.

Results indicate that almost half (46.5%) of the cumulative contribution of demographic and nutrition-related questions to cognitive function was derived from three dietary queries, along with participants’ age (8.8%). The these included consumption of teaspoons of sugar (question 17, 18%), meat, fish, eggs and dairy products (question 31, 13%), and wholegrains (question 16, 6.7%). Based on the top regression coefficients attributed to our dummy variables, greater consumption of added sugar, and sunflower oil was found to be negatively correlated with cognitive function, whereas the regular consumption of wholegrains, green tea/red wine and animal proteins like meat, fish, eggs and dairy products was positively correlated with cognitive function.

### BRAiN scores

We used the regression coefficients obtained from the extended ENR algorithm to compute participants’ BRAiN scores, normalised to the range 0-100. Descriptive statistics for these individual scores are summarised in Table 4. The distribution of BRAiN scores was slightly skewed to the left, as confirmed by both a Kolmogorov-Smirnov test (*p* <2.2e-16), and a D’Agostino K-squared test (s =-0.57, *p* <2.2e-16).

A one-way ANOVA and Tukey post-hoc test revealed significant differences in nutrition scores based on participants’ gender, in that females were found to score significantly higher than males (mean difference: 3.09, *p* <.001)^1^. Additionally, BRAiN scores were found to be significantly higher in individuals older than 65 compared to their younger counterparts (mean difference: 2.47, *p* <0.001).

Spearman’s correlation analyses revealed a positive association between participants’ nutrition and cognitive scores (correlation coefficient: *r* = 0.12, *p* <2.2e-16) and qualitative analyses revealed that both scores steadily declined past a certain point (Figure 9). More specifically, the decrease in CFT scores was found to be statistically significant past the age of 55.29 ± 0.41. By contrast, nutrition scores were initially found to steadily increase, up to age 44.43 ± 0.93. They then remained relatively stable across approximately two decades and steadily decreased past the age of 68.56 ± 1.37. Direct comparisons between the slopes of these two curves after their respective breakpoints using parametric bootstrap (n = 1,000 iterations) yielded a 95% confidence interval for the post-breakpoint slope ranging from –0.64 to –0.60 for CFT scores, and from-0.26 to –0.20 for nutrition scores (observed difference in decline slopes:-0.40). In other words, past these identified breakpoints, CFT scores and nutrition scores were found to decrease by approximately 0.62 and 0.23 points per year, respectively. This divergence in age-related decline between scores was statistically meaningful, as confirmed via nonparametric permutation test (n = 1,000 iterations, *p* = 0.001).

**Figure 9.**
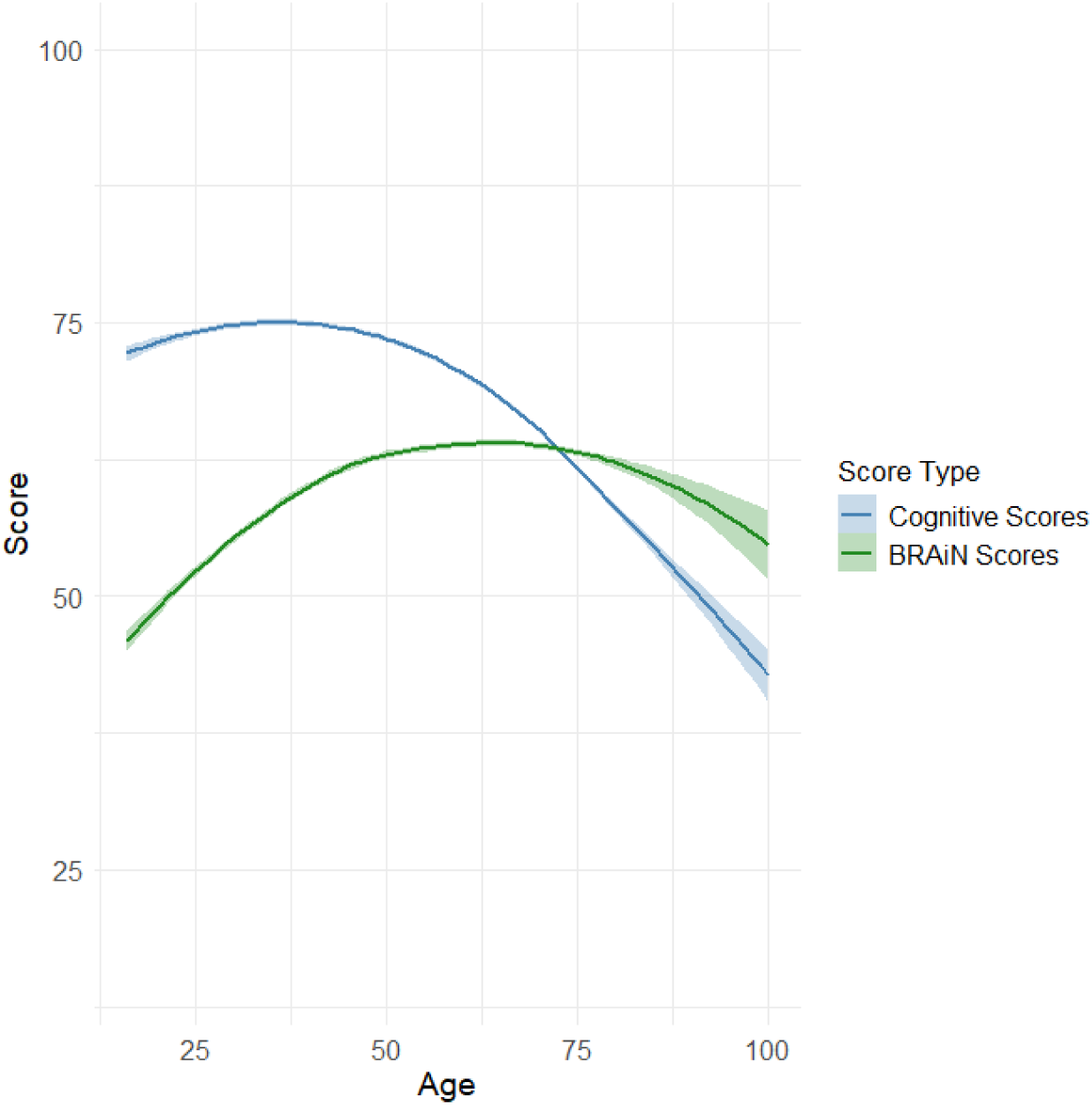
LOESS regression curves for cognitive scores (CFT) and BRAiN scores as a function of participants’ biological age. Both scores were found to decline with increasing age, although these dynamics emerged from different breakpoints: 55.29 ± 0.41 for CFT scores and 68.56 ± 1.37 for BRAiN scores. The rate of decline was also found to be statistically different, whereby, past their identified breakpoints, CFT scores and nutrition scores were found to decrease by approximately 0.62 and 0.23 points per year, respectively.

### Mixed effect ENR model

We built an additional ENR model by introducing interaction terms between age and nutrition-related predictors. This mixed effect model had little effect on MSE in the validation dataset, though it did decrease slightly from 130.97 to 130.70 (R^2^ = 0.16). These results indicate minimal improvements compared to our previous ENR model. In fact, the estimated relative contribution of each interaction term was consistently found to be very small (< 0.07), and the top 25 predictors resulting from this ENR analysis are largely in agreement with those identified in our previous model (Supplementary Figure 5). Taken together, these findings suggest that, although age impacts CFT scores to a meaningful degree, the contributions of nutrition-related dietary choices to participants’ cognition does not vary considerably by age. To further showcase the negligible contribution of interaction terms between age and nutrition-related variables to cognitive function, the cumulative regression coefficients attributed to each nutrition question from the sum of its associated dummy variables and the cumulative regression coefficients of the corresponding interaction terms are presented side-by-side in Supplementary Table 6.

### ENR models by age groups

Lastly, informed by the results of the segmented regression previously described, we split our training and testing datasets into three age groups (16–43-year-olds, 44-68-year-olds and older-than-68) and developed a separate ENR model for each age range. The number of respondents in each group, along with the MSE, R^2^, and the top 10 predictors identified by each regression algorithm are summarised in Tables 5 and 6. Cumulative contributions to CFT score, obtained from the sum of dummy variable coefficients in absolute values, have been highlighted in green and red to indicate the overall positive or negative association with participants’ cognition, respectively. As in previous models, the coefficient attributed to age has been multiplied by a factor of 10 to reflect the effects of this demographic variable on cognitive function by decade. PAge was found to be a significant predictor of cognitive function only starting from the fifth decade of life. In the older group in particular, age was by far the largest predictor, with an estimated contribution to participants’ cognition of 40%, immediately followed by female sex (6.80%), which, importantly, featured among the top 10 predictors only in this cohort. Several nutrition-focused variables consistently appeared within the top 10 predictors regardless of age group (e.g. sugar, animal and vegetable proteins, fried foods), while also being associated with cognitive function in the same (positive or negative) direction, albeit to different extents. Interestingly, however, regular consumption of nuts and seeds was found to be positively associated with CFT in younger (16-43) and older individuals (68+), but not in middle-aged participants (44-68).

**Table 5.** Model statistics and identified predictors from ENR analyses by age group.

| Age group | # of respondents | Baseline MSE | Model MSE | R<sup>2</sup> |
| --- | --- | --- | --- | --- |
| 16-43 | 5880 | 128.27 | 118.92 | 0.07 |
| 44-68 | 16284 | 131.67 | 121.05 | 0.08 |
| 68+ | 6804 | 152.64 | 139.62 | 0.09 |

**Table 6.** Top 10 predictors of cognitive function identified by the ENR algorithm in each age group. Absolute contributions reflect the overall impact each demographic or dietary query has on participants’ cognitive function scores, regardless of the positive or negative effects associated with specific frequencies of consumption (as indicated by the regression coefficients attributed to individual dummy variables).

| Age group | Top 10 predictors | Absolute contribution | Relative weight (%) |
| --- | --- | --- | --- |
| 16-43 | Q17 – Teaspoons of sugar | 6.63 | 18.97 |
|  | Q31 – Animal protein | 3.62 | 10.35 |
|  | Q38 – Sunflower oil | 3.62 | 10.35 |
|  | Q29 – Green tea/Red wine | 3.43 | 9.82 |
|  | Q18 – Fish | 3.36 | 9.60 |
|  | Q23 – Orange/Red vegetables | 3.00 | 8.58 |
|  | Q30 – Vegetable protein | 2.60 | 7.43 |
|  | Q28 – Fried foods | 2.28 | 6.51 |
|  | Q16 – Wholegrains | 1.64 | 4.68 |
|  | Q21 – Nuts and seeds | 1.15 | 3.28 |
| 44-68 | Q17 – Teaspoons of sugar | 4.15 | 17.79 |
|  | Age | 3.45 | 14.79 |
|  | Q31 – Animal protein | 3.27 | 14.04 |
|  | Q24 – Dark leafy vegetables | 2.52 | 10.80 |
|  | Q16 – Wholegrains | 1.60 | 6.87 |
|  | Q38 – Sunflower oil | 1.55 | 6.65 |
|  | Q21 – Nuts and seeds | 1.16 | 4.98 |
|  | Q14 – Sugary snacks/drinks | 1.05 | 4.52 |
|  | Q29 – Green tea/Red wine | 1.03 | 4.40 |
|  | Q30 – Vegetable protein | 0.86 | 3.69 |
| 68+ | Age | 6.76 | 40.00 |
|  | Gender (Female) | 1.15 | 6.80 |
|  | Q20 – Nuts and seeds | 1.10 | 6.50 |
|  | Q17 – Teaspoons of sugar | 1.03 | 5.37 |
|  | Q34 – Coffee | 0.91 | 4.89 |
|  | Q28 – Fried foods | 0.83 | 3.60 |
|  | Q31 – Animal protein | 0.82 | 3.39 |
|  | Q30 – Vegetable protein | 0.61 | 2.70 |
|  | Q22 – Berries | 0.57 | 2.69 |
|  | Gender (Male) | 0.46 | 2.54 |

## Discussion

The present study used a large dataset of self-reported measures to explore potential associations between dietary intake and performance on the Food for the Brain online CFT. The methodological approach chosen for this study was motivated by the desire to identify nutrition-related predictors of cognitive function without any pre-existing assumptions regarding either the relative impact of each dietary variable on participants’ cognition, or the mathematical relationship between the frequency of consumption of specific foods or food groups and cognitive scores. The implementation of our ENR algorithm, along with the use of dummy variables, provided estimates for the relative contribution of each dietary metric, as well as fine-grained predictions of the positive or negative associations related to different patterns of consumption (e.g. different number of servings per week) on CFT. By considering the entire set of nutritional variables at our disposal, we therefore investigated significant associations between participants’ eating habits and cognitive function in the context of their entire food matrix rather than teasing apart isolated relationships.

Targeted analyses comparing a data-driven score (i.e. MIND* ENR) and MIND* Linear score suggested that our statistical approach yielded greater accuracy in the prediction of participants’ cognitive function, as demonstrated by significantly greater correlation with CFT scores in the validation dataset. A closer look at the regression coefficients attributed to each dummy variable revealed that assumptions of linearity between all potential answers for the same dietary question may result in lower predictive accuracy. Results further revealed that greater consumption of sugar-based snacks or drinks is associated with lower CFT scores, while greater consumption of wholegrains, vegetables, vegetable proteins, and nuts and seeds is associated with higher CFT scores. Surprisingly, higher intakes of berries and fish showed an inverse relationship with CFT score, while the consumption of fried and fast foods was positively associated with CFT score. These counterintuitive findings may have been driven by demographic and socio-economic factors, as suggested by the distribution of participants’ responses by age group (Figures 3-5). Younger individuals, who tend to outperform their older counterparts on cognitive tests, consume convenient, more affordable food items such as fried and fast foods more regularly, and incorporate relatively more expensive products such as berries and fish more sporadically. As socio-economic determinants were not accounted for in the current work, future investigations and/or survey iterations including additional ecological variables may help elucidate these intricate dynamics. Follow-up score propensity analyses on berries and fish consumptions including respondents’ demographic characteristics and supplementation strategies further revealed that in fact higher intakes of berries are associated with higher CFT scores on average, while the association between higher intakes of fish and impoverished cognitive function persisted. While it is reasonable to suggest that these isolated findings may reflect reverse causation, in that people with declining cognition (who are typically older) might be increasing their fish intake and/or supplement intake because they believe them to be neuroprotective, this hypothesis could not be tested directly using the data at our disposal.

The subsequent introduction of a wider range of nutrition-related variables, using ENR to analyze all available nutrition-related Food for the Brain questions and compute a BRAiN score for each participant, allowed us to improve model performance to a significant extent, as indicated by a lower MSE and a greater proportion of the variance being explained by the model itself. These findings reinforce the idea that participants’ entire dietary pattern should be considered when evaluating the relationship between nutrition and cognitive function, above and beyond the consumption of selected foods and food groups that have been previously shown to affect overall cognition, either in the positive or in the negative direction.

Results from this extended regression analysis indicated that the regular consumption of both added sugar and sunflower oil (the most common refined oil in the UK where the majority of participants live) is negatively associated with cognitive function. By contrast, positive associations were observed when consuming animal and vegetable proteins, as well as vegetables, wholegrains, nuts and seeds. These findings corroborate the conclusions of similar lines of research showing the beneficial link between diets centred around whole foods and cognition, including both observational studies and randomized controlled trials^23, 24^. Nutrition scores were also found to be higher in females compared to males, as well as in older (> 65 years) compared to younger participants, highlighting non-negligible differences in nutrition practices based on both gender and age. CFT scores were also higher in females but lower in older participants. Both cognitive and nutrition scores were found to decline as age increased, although at different rates. More precisely, we observed a sharp downward inflection in CFT scores starting at approximately 55 years of age. By contrast, nutrition scores showed an initial increase, up to age 44, followed by a sustained plateau and a statistically significant decline later in life, shortly past the 68 years’ mark. This intriguing disparity may speak to the chronic nature of age-related cognitive decline, in that the physiological and metabolic foundation for the development of Alzheimer’s dementia may be laid well before the appearance of the first noticeable symptoms^25, 26^, and the collection and analysis of longitudinal data will be crucial to corroborate this working hypothesis. By contrast, nutrition habits may be expected to improve as individuals approach the central decades of their life due to greater financial stability and access to nutritious food, as well as increased appreciation for sound nutritional practices and their direct consequences on health and well-being. However, the risk of malnutrition and dietary inadequacies significantly increases in older age due to a combination of physical, physiological, societal and environmental factors deeply affecting an individual’s relationship with food^21, 27^, which may explain the ultimate decline in nutrition scores we detected in the later decades of life.

Perhaps surprisingly, mixed effect regression analyses including interaction terms between nutrition variables and age did not improve our model performance to an appreciable degree. Along the same lines, when looking at dietary habits within specific age groups, the greatest predictors of cognitive functions were found to be largely the same across the board. How exactly age and nutrition interact in driving cognitive outcomes in the later decades of life is still not entirely understood, although our data suggest that even nutritionally sound dietary practices are unlikely to completely counteract age-related physiological mechanisms that inevitably affect cognitive function.

Lastly, our analyses demonstrated a small, negative correlation between participants’ MIND* Linear scores and cognitive scores. This key finding sheds new light on the validity and real-life applicability of current nutritional approaches to dementia prevention, while also calling for the recasting of current dietary guidelines based on the evidence provided by relevant population data^28^.

For example, the MIND diet was explicitly designed in the setting of Western-style diets, raising the question of how well this index would fare when applied to Asian, African, or indigenous populations where culinary traditions are remarkably distinct^29^. While formal evaluations of the MIND diet, or some adaptation thereof, are slowly starting to emerge (e.g., in Pakistan^30^ and China^31^), it is reasonable to argue that the questions included in the original MIND model may not be appropriate to adequately capture the complex interplay between nutritional habits and cognition across the globe.

Irrespective of these methodological and cultural limitations, the effectiveness of the MIND dietary index in its current formulation will likely be reduced or even become outdated at some point, as dietary trends naturally shift over time in response to environmental determinants like changes in the food supply chain (e.g., improved availability and access to certain foods, introduction of innovative food products, phase-out of specific food items, etc.), improved socio-economic status and technological development, along with better understanding of the impact dietary choices have on planetary health, physical health and wellbeing. The validity and reliability of this nutritional approach should therefore be regularly re-assessed, and dietary guidelines should be adjusted to reflect any changes that may have been introduced in common dietary practices.

Our proposed mathematical formulation presents the unique advantage of being largely dynamic and adaptable in nature, allowing for ad-hoc and timely adjustments to current nutrition recommendations based on ongoing dietary trends, as well as tuning to specific cultures across geographic areas of the globe. More specifically, we envisage that the statistical model described in this work could be applied to large population datasets at regular intervals (e.g. annually). This could even be done on a continuous, ongoing basis if there were sufficient uptake of recurring assessments longitudinally. The predictions obtained from each new model application could then be used to (i) refine the formal definition of nutrition scores, (ii) assess the adequacy of the nutrition guidelines promoted at each point in time against the incidence or progression of age-related cognitive decline, and (iii) inform global health authorities on necessary updates to dietary recommendations targeting dementia prevention.

While the CFT used here is still relatively underexploited, it is already a large epidemiological dataset, and several promising avenues of research could be pursued if the test was administered more systematically and on a larger, global scale. Firstly, as more data become available from more countries across the globe, the statistical power of our model will inevitably increase, thus leading to more accurate predictions. Secondly, if respondents continue to take the cognitive function test every year, multiple survey entries from the same subjects could be incorporated in the present model to help track each participant’s cognitive status longitudinally, along with concomitant changes in behavioural and lifestyle factors. For example, by selecting cohorts of relatively young or middle-aged participants on the one hand, and groups of older individuals with similar lifestyle habits on the other, the model could generate predictions on the prospective cognitive health of the younger cohort based on cognitive outcomes observed in the older group. Over time, these predictions could then be validated with real, longitudinal data from the younger cohort and compared to current estimates of dementia incidence so that adjustments could be made to the model that progressively improve predictive accuracy.

Finally, and perhaps most importantly, with the help of healthcare providers and global health authorities, the completion of the Food for the Brain Cognitive Function Test, along with a standardised medical check-up and comprehensive blood panel, could be promoted at the population level as routine cognitive assessments. Given the large variability in both nutritional and lifestyle practices within our society, based on factors such as geographical location, cultural traits and economic status^32–34^, each country could be responsible for the setup of its own cognitive health initiative and the collection of relevant data to help shape national guidelines that better apply to their citizens.

To summarise, this study presents several strengths, primarily due to the use of large sets of population data and the implementation of a data-driven statistical algorithm that is not constrained by a priori assumptions on the relationship between potential predictors and outcome variables. Nonetheless, given the unavoidable practical determinants related to data collection, our findings should be interpreted with caution and numerous study limitations warrant discussion.

First of all, as is often the case with health and lifestyle questionnaires^35^, common issues of self-reported measurements such as the risk of respondent or recall biases must be considered^36^. Food frequency questionnaires are notoriously prone to these errors.^37, 38^ Certain questions included in the survey could be reformulated to increase accuracy, reduce participants’ imprecision or uncertainty, and minimize the occurrence of errors when completing the survey. For instance, separate questions could have been asked regarding participants’ consumption of animal proteins, such as red and white meats, fish, eggs, and dairy products. Given that all these items are currently bundled together within the same question, it is impossible to disentangle the relevant contributions of each protein type to cognitive function. In fact, due to their high content of omega-3 fatty acids and choline, respectively, fish and eggs could be expected to exert beneficial effects on overall cognition^39–41^, whereas the relatively high amounts of saturated fats found in certain red meats and dairy products may encourage more moderate intakes to decrease risk of cognitive delcine^42^. At present, results from our analyses indicate that the regular consumption of these foods taken together is positively correlated with cognitive function, yet it remains unclear if this result is driven by an overall neuroprotective benefit related to all food items included in the question, or rather from the positive net balance of their combined contributions, whereby selected nutrients found in certain foods only (e.g. omega-3s, choline)^43, 44^ may offset the potential harm caused by selected compounds found in other (e.g. saturated fats)^45^, that may raise ApoB and therefore increase dementia risk through higher risk of cardiovascular disease. Finally, as our goal was primarily focused on prediction, the best coefficients for predicting CFT do not necessarily map onto dietary recommendations for improving cognitive function through diet, and the associations noted cannot be assumed to have causal effects.

In our attempt to improve the design of subjective assessment methods to investigate eating behaviours on a global scale, it is worth pointing out that the remarkable diversity of current dietary patterns, driven by geographical, cultural and socio-economic factors alike, is inherently difficult to represent using a single, standardised instrument. As in the case of the MIND diet index, the FFB survey was also shaped around eating behaviours typically observed in the Western world and data employed in this study mostly came from a few of these predominantly English-speaking countries, which may have skewed our results based on the dietary trends currently followed in these regions of the world. Whether our findings would be consistently replicated in culturally different cohorts remains to be determined. Moving forward, as more data are hopefully collected from various areas across the globe, it will be crucial to routinely assess whether adjustments to the survey content or formulation are required, both to reflect potential shifts in dietary trends over time, and to ensure that the reliability and validity of this measure is preserved regardless of societal and cultural determinants. This systematic re-evaluation process would be particularly relevant to developing countries and areas of the world where the food environment is rapidly shifting away from long-standing culinary traditions and towards more processed foods, fried foods, and sugary foods and drinks. Finally, in recent years, novel pharmacological agents to tackle the current obesity epidemic (e.g. GLP-1 agonists like semaglutide and tirzepatide) have started to emerge. While still in their infancy, these tools are progressively taking centre stage and have the potential to modify our individual and societal behaviours towards dietary practices to a significant extent. The increasingly popular use of these drugs adds another level of complexity to the dynamic interrelationship between diet and cognition, thus reinforcing the idea that reliable statistical models of cognitive function must be dynamical in nature to yield reliable predictions while accounting for all these interacting components.

The more we understand about the relationship between cognitive function and modifiable risk factors, the more questions could be added or edited to investigate a particular variable or lifestyle behaviour in greater detail. It is easy to appreciate that our quest is mostly limited by the type of information we can gather from the current survey and its level of detail. Although longitudinal comparisons would be harder to perform if the survey were to be modified in the future, small edits to certain fundamental questions could increase the accuracy of participants’ responses to a significant degree and might therefore be worthy of consideration. Related to this point, our definition of MIND* scores did not perfectly follow the original MIND model due to the absence or different formulation of a few relevant questions. Once again, this issue could be taken into account if changes were to be made to the current survey to allow for better statistical comparisons between dietary indices in future survey iterations. As a final note, we stress that dietary practices certainly do not affect cognitive function in isolation and several other non-modifiable (e.g. genetics) and modifiable risk factors (e.g. sleep, physical activity, social behaviours, stress) undeniably play a prominent role in driving cognitive outcomes in older age^46,47^. However, exploring the even more complex interplay among the numerous dimensions of human health and cognitive ageing was beyond the scope of this study.

## Conclusion

The present study investigated the impact of self-reported dietary practices on predicted cognitive function using an elastic net regression model. This statistical approach informed the definition of data-driven BRAiN scores that more closely correlated with respondents’ overall cognition when compared to traditional MIND scores. Our analyses further revealed significant contributions to respondents’ cognitive function scores deriving from the regular consumption of selected foods. More precisely, animal proteins and wholegrains were found to be positively associated with participants’ cognition, whereas the opposite relationship emerged with respect to added sugar and sunflower oil, which associated negatively with cognitive function scores. Paired with a more widespread employment of the Food for the Brain Cognitive Function Test and lifestyle questionnaire, the adoption of this novel method of enquiry could be used to track shifts in nutritional trends over time, identify potential dietary gaps within the population, and promptly adjust dietary recommendations and guidelines to better address nutrient inadequacies that may negatively affect cognitive health, especially in older age.

## Author Contributions

Conceptualisation, F.C. and T.R.W.; methodology, F.C. and T.R.W.; investigation, F.C., K.G., A.S. and T.R.W.; writing—original draft preparation, F.C.; writing—review and editing, F.C., K.G., A.S. and T.R.W.; supervision, T.R.W.; funding acquisition, T.R.W. All authors have read and agreed to the published version of the manuscript.

## Statements and Declarations

### Ethical considerations

the University of Washington Institutional Review Board confirmed that this work did not constitute human subjects research and ethical approval was not necessary for the conduct of the present study.

## Funding

This research received no external funding.

### Conflicts of Interest

T.R.W. is a paid scientific advisor for Hintsa Performance, Thriva LLC., and Better Brain LLC.

## Data Availability

All data produced in the present study are available upon reasonable request to the authors

## Footnotes

1 Participants’ cognitive function test scores were also found to be significantly higher in females than males (mean difference: 1.38, p<2.2e-16).

## Notes

### Funding Statement

This study did not receive any funding

### Author Declarations

The University of Washington Institutional Review Board confirmed that this work did not constitute human subjects research and ethical approval was not necessary for the conduct of the present study.

## REFERENCES

1. Livingston G, Huntley J, Liu KY, et al. Dementia prevention, intervention, and care: 2024 report of the Lancet standing Commission. The Lancet 2024; 404: 572–628. DOI: 10.1016/S0140-6736(24)01296-0.

2. Nichols E, Steinmetz JD, Vollset SE, et al. Estimation of the global prevalence of dementia in 2019 and forecasted prevalence in 2050: an analysis for the Global Burden of Disease Study 2019. The Lancet Public Health 2022; 7: e105–e125.

3. Hendriks S, Peetoom K, Bakker C, et al. Global incidence of young-onset dementia: A systematic review and meta-analysis. Alzheimer’s & Dementia 2023; 19: 831–843. DOI: 10.1002/alz.12695.

4. Seifert I, Wiegelmann H, Lenart-Bugla M, et al. Mapping the complexity of dementia: factors influencing cognitive function at the onset of dementia. BMC Geriatrics 2022; 22: 507. DOI: 10.1186/s12877-022-02955-2.

5. Gao Y, Zhang Z, Song J, et al. Combined healthy lifestyle behaviours and incident dementia: A systematic review and dose–response meta-analysis of cohort studies. International Journal of Nursing Studies 2024; 156: 104781. DOI: 10.1016/j.ijnurstu.2024.104781.

6. Hillari L, Frank P and Cadar D. Systemic inflammation, lifestyle behaviours and dementia: A 10-year follow-up investigation. *Brain, Behavior*, & Immunity - Health 2024; 38: 100776. DOI: 10.1016/j.bbih.2024.100776.

7. Baker LD, Espeland MA, Whitmer RA, et al. Structured vs Self-Guided Multidomain Lifestyle Interventions for Global Cognitive Function: The US POINTER Randomized Clinical Trial. JAMA 2025. DOI: 10.1001/jama.2025.12923.

8. Boa Sorte Silva NC, Barha CK, Erickson KI, et al. Physical exercise, cognition, and brain health in aging. Trends in Neurosciences 2024; 47: 402–417. DOI: 10.1016/j.tins.2024.04.004.

9. Zhang Y, Chen S-D, Deng Y-T, et al. Identifying modifiable factors and their joint effect on dementia risk in the UK Biobank. Nature Human Behaviour 2023; 7: 1185–1195. DOI: 10.1038/s41562-023-01585-x.

10. Morris MC, Tangney CC, Wang Y, et al. MIND diet slows cognitive decline with aging. Alzheimer’s & Dementia 2015; 11: 1015–1022. DOI: 10.1016/j.jalz.2015.04.011.

11. Thomas A, Lefèvre-Arbogast S, Féart C, et al. Association of a MIND Diet with Brain Structure and Dementia in a French Population. J Prev Alzheimers Dis 2022; 9: 655–664. DOI: 10.14283/jpad.2022.67.

12. McEvoy CT, Guyer H, Langa KM, et al. Neuroprotective Diets Are Associated with Better Cognitive Function: The Health and Retirement Study. Journal of the American Geriatrics Society 2017; 65: 1857–1862. DOI: 10.1111/jgs.14922.

13. Kheirouri S and Alizadeh M. MIND diet and cognitive performance in older adults: a systematic review. Critical Reviews in Food Science and Nutrition 2022; 62: 8059–8077. DOI: 10.1080/10408398.2021.1925220.

14. Aggarwal NT, Dhana K, Agarwal P, et al. The role of dietary fatty acids intake in the association between cortical thickness and global cognitive function: The MIND trial. Alzheimer’s & Dementia 2020; 16: e045260. DOI: 10.1002/alz.045260.

15. Hosking DE, Eramudugolla R, Cherbuin N, et al. MIND not Mediterranean diet related to 12-year incidence of cognitive impairment in an Australian longitudinal cohort study. Alzheimer’s & Dementia 2019; 15: 581–589. DOI: 10.1016/j.jalz.2018.12.011.

16. Devranis P, Vassilopoulou Ε, Tsironis V, et al. Mediterranean diet, ketogenic diet or MIND diet for aging populations with cognitive decline: a systematic review. Life 2023; 13: 173.

17. Barnes LL, Dhana K, Liu X, et al. Trial of the MIND diet for prevention of cognitive decline in older persons. New England Journal of Medicine 2023; 389: 602–611.

18. Trustram Eve C and de Jager CA. Piloting and validation of a novel self-administered online cognitive screening tool in normal older persons: The Cognitive Function Test. International journal of geriatric psychiatry 2014; 29: 198–206.

19. Friedman JH, Hastie T and Tibshirani R. Regularization Paths for Generalized Linear Models via Coordinate Descent. Journal of Statistical Software 2010; 33: 1–22. DOI: 10.18637/jss.v033.i01.

20. Zou H and Hastie T. Regularization and variable selection via the elastic net. Journal of the Royal Statistical Society Series B: Statistical Methodology 2005; 67: 301–320.

21. Amarya S, Singh K and Sabharwal M. Changes during aging and their association with malnutrition. Journal of Clinical Gerontology and Geriatrics 2015; 6: 78–84. DOI: 10.1016/j.jcgg.2015.05.003.

22. Grady C. The cognitive neuroscience of ageing. Nature Reviews Neuroscience 2012; 13: 491–505. DOI: 10.1038/nrn3256.

23. Jiwani R, Robbins R, Neri A, et al. Effect of Dietary Intake Through Whole Foods on Cognitive Function: Review of Randomized Controlled Trials. Current Nutrition Reports 2022; 11: 146–160. DOI: 10.1007/s13668-022-00412-5.

24. Townsend RF, Logan D, O’Neill RF, et al. Whole Dietary Patterns, Cognitive Decline and Cognitive Disorders: A Systematic Review of Prospective and Intervention Studies. Nutrients 2023; 15: 333.

25. Antal BB, van Nieuwenhuizen H, Chesebro AG, et al. Brain aging shows nonlinear transitions, suggesting a midlife “critical window” for metabolic intervention. Proceedings of the National Academy of Sciences 2025; 122: e2416433122. DOI: doi:10.1073/pnas.2416433122.

26. Robinson L, Tang E and Taylor J-P. Dementia: timely diagnosis and early intervention. BMJ: British Medical Journal 2015; 350: h3029. DOI: 10.1136/bmj.h3029.

27. Agarwal E, Miller M, Yaxley A, et al. Malnutrition in the elderly: A narrative review. Maturitas 2013; 76: 296–302. DOI: 10.1016/j.maturitas.2013.07.013.

28. European Commission: Directorate-General for Health and Food Safety and ICF Consulting Services. Review of scientific evidence and policies on nutrition and physical activity – A comprehensive review of the scientific evidence about the source of calories consumed and types and frequency of physical activity among Europeans. 2018. Publications Office of the European Union.

29. Timlin D, McCormack JM, Kerr M, et al. The MIND diet, cognitive function, and well-being among healthy adults at midlife: a randomised feasibility trial. BMC Nutrition 2025; 11: 59. DOI: 10.1186/s40795-025-01020-6.

30. Islam B, Li T, Ibrahim TI, et al. The relationship between levels of physical activity, adherence to the MIND diet, and cognitive impairment in adults aged 65 years or older in Pakistan. Journal of Alzheimer’s Disease Reports 2025; 9: 25424823241290132.

31. Lu L, Cai S, Xiao Q, et al. The association between Chinese adapted MIND diet and cognitive function in Chinese middle-aged and older adults: results from the Chinese Square Dance Cohort. European Journal of Nutrition 2025; 64: 22.

32. Bennett G, Bardon LA and Gibney ER. A Comparison of Dietary Patterns and Factors Influencing Food Choice among Ethnic Groups Living in One Locality: A Systematic Review. Nutrients 2022; 14: 941.

33. De Vito R, Stephenson B, Sotres-Alvarez D, et al. Identifying and characterizing shared and ethnic background site-specific dietary patterns in the Hispanic Community Health Study/Study of Latinos (HCHS/SOL). Nutrition Journal 2025; 24: 71. DOI: 10.1186/s12937-025-01138-0.

34. Desbouys L, Méjean C, De Henauw S, et al. Socio-economic and cultural disparities in diet among adolescents and young adults: a systematic review. Public Health Nutrition 2020; 23: 843–860. 2019/08/30. DOI: 10.1017/S1368980019002362.

35. Wang Q, Wang Y, Xie X, et al. The measurement properties of existing lifestyle assessment tools are suboptimal: a systematic review. Journal of Public Health 2023. DOI: 10.1007/s10389-023-02102-0.

36. Miller TM, Abdel-Maksoud MF, Crane LA, et al. Effects of social approval bias on self-reported fruit and vegetable consumption: a randomized controlled trial. Nutrition Journal 2008; 7: 18. DOI: 10.1186/1475-2891-7-18.

37. Mertens E, Kuijsten A, Geleijnse JM, et al. FFQ versus repeated 24-h recalls for estimating diet-related environmental impact. Nutrition Journal 2019; 18: 2. DOI: 10.1186/s12937-018-0425-z.

38. Popoola AA, Frediani JK, Hartman TJ, et al. Mitigating underreported error in food frequency questionnaire data using a supervised machine learning method and error adjustment algorithm. BMC Medical Informatics and Decision Making 2023; 23: 178. DOI: 10.1186/s12911-023-02262-9.

39. Sultan N, Cheng E, McMahon C, et al. The impact of egg consumption on cognitive function: a systematic literature review. Proceedings of the Nutrition Society 2024; 83: E181. 2024/05/07. DOI: 10.1017/S002966512400199X.

40. Yamashita S, Kawada N, Wang W, et al. Effects of egg yolk choline intake on cognitive functions and plasma choline levels in healthy middle-aged and older Japanese: a randomized double-blinded placebo-controlled parallel-group study. Lipids in Health and Disease 2023; 22: 75. DOI: 10.1186/s12944-023-01844-w.

41. Marcos L and Javier G. Cognition and Omega-3 Fatty Acids-A Narrative Review of the Literature. Biomedical Journal of Scientific & Technical Research 2021; 40: 32493–32499.

42. Butler MJ, Mackey-Alfonso SE, Massa N, et al. Dietary fatty acids differentially impact phagocytosis, inflammatory gene expression, and mitochondrial respiration in microglial and neuronal cell models. Frontiers in Cellular Neuroscience 2023; 17: 1227241.

43. Wallace TC. A comprehensive review of eggs, choline, and lutein on cognition across the life-span. Journal of the American College of Nutrition 2018; 37: 269–285.

44. Sherzai AZ, Sherzai AN and Sherzai D. A Systematic Review of Omega-3 Consumption and Neuroprotective Cognitive Outcomes. American Journal of Lifestyle Medicine 2020; 0: 15598276221117102. DOI: 10.1177/15598276221117102.

45. Okereke OI, Rosner BA, Kim DH, et al. Dietary fat types and 4-year cognitive change in community-dwelling older women. Ann Neurol 2012; 72: 124–134. 20120518. DOI: 10.1002/ana.23593.

46. Puri S, Shaheen M and Grover B. Nutrition and cognitive health: A life course approach. Frontiers in public health 2023; 11: 1023907.

47. Glans I, Nägga K, Gustavsson A-M, et al. Associations of modifiable and non-modifiable risk factors with cognitive functions – a prospective, population-based, 17 years follow-up study of 3,229 individuals. Alzheimer’s Research & Therapy 2024; 16: 135. DOI: 10.1186/s13195-024-01497-6.

