## Supplementary Material for "Bias-free Regression Analysis in Nutrition (BRAiN) Index: A Predictive Model of Cognitive Function"

<u>**Contents:**</u>

**S1. Supplementary Figure 1**: Flowchart of data cleaning process.

**S2. Nutrition-focused questions from the health and lifestyle questionnaire and feature re-engineering.**

**S3. Supplementary Figure 2**: Distribution of participants' responses to the nutrition-focused questions from the health and lifestyle survey.

**S4. Supplementary Table 2:** Question features re-engineering**.**

**S5. Supplementary Figure 3.** Distributions of participants' responses to the 10 queries selected for the calculation of individual MIND* scores.

**S6. Comparison between elastic net regression and linear regression models performance.**

**S7. Supplementary Table 5**: Survey questions and linear scales used to calculate the MIND* Linear score following the original MIND questions.

**S8. Supplementary Figure 5**: World map of study participants providing cultural background information.

**S9. Distribution of responses to questions 18, 22 and 28 by education group.**

**S10. ENR Regression coefficients including interaction terms with age.**

**S1. Supplementary Figure 1**: Flowchart of data cleaning process.

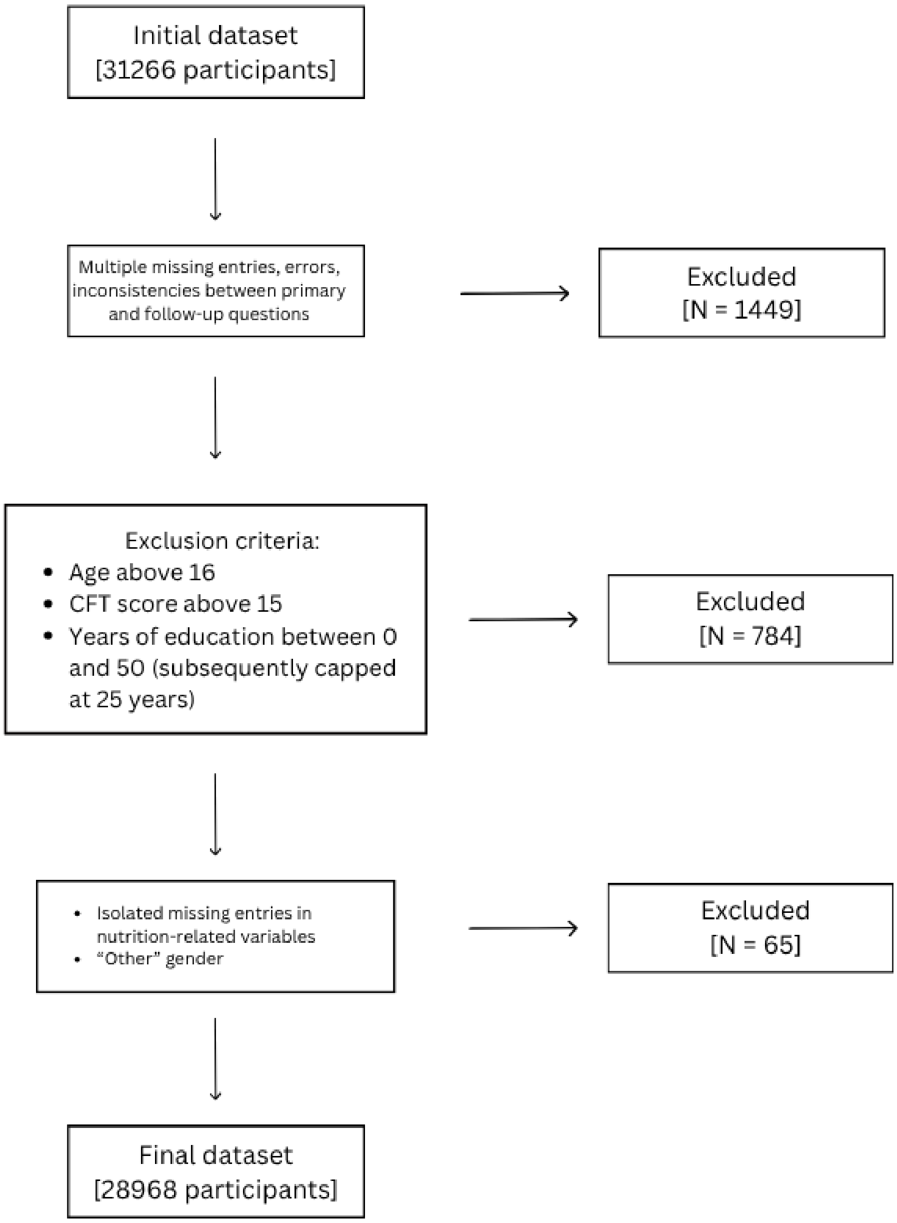

### S2. Nutrition-focused questions from the health and lifestyle questionnaire and feature re-engineering

The nutrition-focused questions from the Food for the Brain health and lifestyle questionnaire are presented in Supplementary Table 1, alongside their five possible answers. A closer look at the distributions of participants' responses to these questions revealed that some of these distributions were heavily skewed due to certain categories being endorsed by a small proportion of respondents only (see Supplementary Figure 2). Consequently, to avoid potential biases or statistical misrepresentations driven by these lower percentages, prior to data analysis, participants answers were carefully regrouped into three levels to ensure each category was endorsed by at least 10% of our respondents. The updated categories resulting from the recoding process are presented in Supplementary Table 2.

**Supplementary Table 1**: Original nutrition-focused questions from the health and lifestyle questionnaire.

| Question | Possible answers | | | | |
|---|---|---|---|---|---|
| **Q14:** How many sugar-based snacks or drinks (biscuits, cakes, chocolate, sweets, fizzy drinks, fruit juices) do you consume each day? | None/Rarely | 1 a day or less | 2 a day | 3 a day | 3+ a day |
| **Q15:** How many times a day do you eat white rice, white bread, flour or other refined foods? | None/Rarely | 1 a day or less | 2 a day | 3 a day | 3+ a day |
| **Q16:** How many times a week do you eat wholegrains e.g. Brown rice, oats, wholegrain bread, wholewheat pasta? | None | 1 a week or less | 2-3 a week | 3-7 a week | Every day |
| **Q17**: How many teaspoons (or equivalent) of sugar do you add to food or drinks each day? | None/Rarely | 1 a day or less | 2-4 a day | 4-7 a day | 7+ a day |
| **Q18:** How many times a week do you eat fish (of any kind)? | None | Less than 1 a week | 1 a week | 2 a week | 3+ a week |
| **Q19:** How many times a week do you eat fresh oily fish (e.g. salmon, mackerel, sardines, herring)? | None | Less than 1 a week | 1 a week | 2 a week | 3+ a week |
| **Q20:** How frequently do you eat raw walnuts, pecans, macadamia, chia or flax seeds? (a handful or small cup) | Never | Rarely/Occasionally | 2-3 a week | 3-5 a week | At least 1 a day |
| **Q21:** How many servings of fresh fruit do you have a day? | Never/Rarely | Less than 1 a day | 1 a day | 2-3 a day | 3+ a day |

| | | | | | |
|---|---|---|---|---|---|
| **Q22:** How many servings of berries, cherries, plums, pomegranate or their juice do you have a day? | Never/Rarely | 1-2 a week | 4 or more a week | Most days | 2 or more a day |
| **Q23:** How many servings of orange or red vegetables do you have a week (e.g. carrot, beetroot, sweet potato, squash, peppers? | None | 1 a week or less | 2-3 a week | 3-4 a week | 5+ a week |
| **Q24:** How many servings of green leafy vegetables do you have a week? (e.g. kale, spinach, lettuce, watercress, rocket, cabbage, greens) | None | 1 a week or less | 2-3 a week | 3-4 a week | 5+ a week |
| **Q27:** How many times a week do you eat dark green or cruciferous vegetables? (e.g. broccoli, cabbage, cauliflower, brussels sprouts)? | Never/Rarely | 1 a week or less | 2-3 a week | 3-6 a week | Most days |
| **Q28:** How many times a week do you eat fried, deep fried or browned foods including crisps, chips and fried takeaway food? | Never/Rarely | 1 a week or less | 2-3 a week | 3-6 a week | Most days |
| **Q29:** How many green or herbal teas or small glasses (175mls) of red wine do you drink a week? | None/Rarely | 1 to 3 | 4 to 6 | 7 to 11 | 12 or more |
| **Q30:** How many times a day do you eat vegetable protein? (e.g. beans, lentils, tofu, quinoa, peas, corn) | None/Rarely | 1 or 2 a week | Every other day | Once a day | 2+ a day |
| **Q31:** How many times a week do you eat a serving of meat, fish, eggs, cheese or dairy products? | None/Rarely | 1 or 2 a week | Every other day | Once a day | 2+ a day |
| **Q32:** How many slices of bread, biscuits, or servings of pasta or pizza do you have each day? | None/Rarely | 1 a day or less | 2-3 a day | 4-6 a day | 6+ a day |
| **Q34:** How many coffees (excluding decaf) do you have a day (a double expresso counts as two)? | None/Rarely | 1 a day or less | 2-3 a day | 4-6 a day | 6+ a day |
| **Q38:** How often do you use sunflower oil in cooking, dressings, etc.? | Never | Rarely | Sometimes | Frequently | Often |
| **Q39:** How many eggs do you eat each week? | None | 1 to 2 | 3 to 4 | 5 to 6 | More than 6 |
| **Q40:** Are you vegan (completely avoid animal products including meat, fish, eggs and dairy)? | No | | | | Yes |

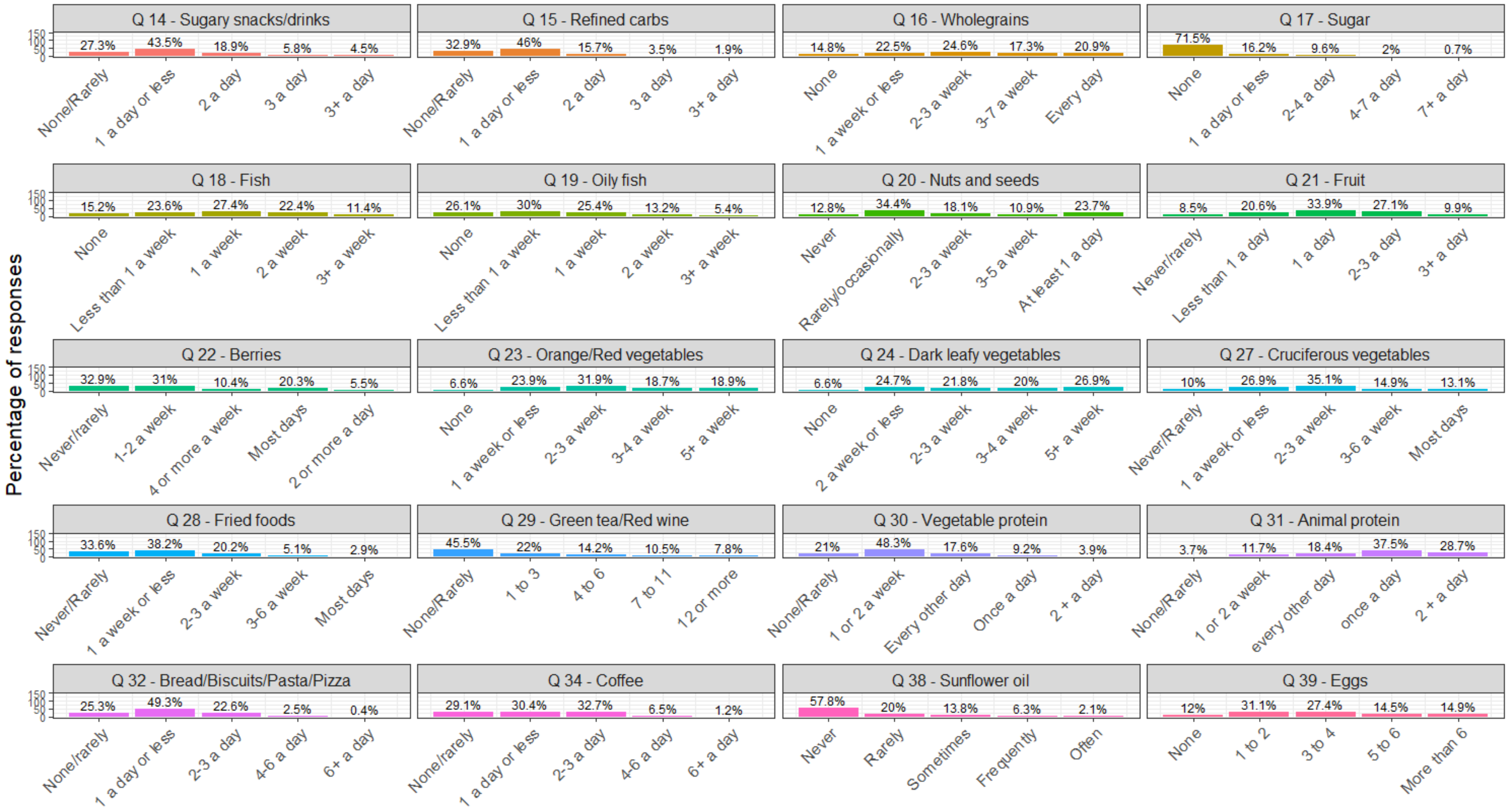


**S3. Supplementary Figure 2**: Distribution of participants' responses to the nutrition-focused questions from the health and lifestyle survey.

**S4. Supplementary Table 2**: Question features re-engineering.

| Question | Possible answers | | |
|---|---|---|---|
| **Q14:** How many sugar-based snacks or drinks (biscuits, cakes, chocolate, sweets, fizzy drinks, fruit juices) do you consume each day? | None/Rarely | 1 a day or less | 2+ a day |
| **Q15/32:** How many times a day do you eat refined carbohydrates? | None/Rarely | 1 a day or less | 2+ a day |
| **Q16:** How many times a week do you eat wholegrains e.g. Brown rice, oats, wholegrain bread, wholewheat pasta? | None | 1 a week or less | 2-3 a week |
| **Q17**: How many teaspoons (or equivalent) of sugar do you add to food or drinks each day? | None/Rarely | 1 a day or less | 2+ a day |
| **Q18/19:** How many times a week do you eat fish (of any kind)? | None/Rarely | 1-2 a week | 3+ a week |
| **Q20:** How frequently do you eat raw walnuts, pecans, macadamia, chia or flax seeds? (a handful or small cup) | Never/Rarely/ Occasionally | 2-3 a week | 3+ a week |
| **Q21:** How many servings of fresh fruit do you have a day? | Less than 1 a day | 1 a day | 2+ a day |
| **Q22:** How many servings of berries, cherries, plums, pomegranate or their juice do you have a day? | Never/Rarely | 1-2 a week | Most days or more |
| **Q23:** How many servings of orange or red vegetables do you have a week (e.g. carrot, beetroot, sweet potato, squash, peppers? | 1 a week or less | 2-4 a week | 2-3 a week |
| **Q24/27:** How many servings of green leafy or cruciferous vegetables do you have a week? | 2 a week or less | 3-4 a week | 5+ a week |
| **Q28:** How many times a week do you eat fried, deep fried or browned foods including crisps, chips and fried takeaway food? | Never/Rarely | 1 a week or less | 2+ a week |
| **Q29:** How many green or herbal teas or small glasses (175mls) of red wine do you drink a week? | None/Rarely | Regularly | 7+ a week |
| **Q30:** How many times a day do you eat vegetable protein? (e.g. beans, lentils, tofu, quinoa, peas, corn) | None/Rarely | 1 or 2 a week | 3+ a week |
| **Q31:** How many times a week do you eat a serving of meat, fish, eggs, cheese or dairy products? | None/Rarely/ Infrequently | Every other day | Daily |
| **Q34:** How many coffees (excluding decaf) do you have a day (a double expresso counts as two)? | None/Rarely | 1 a day or less | 2+ a day |
| **Q38:** How often do you use sunflower oil in cooking, dressings, etc.? | Never | Rarely | Regularly |
| **Q39:** How many eggs do you eat each week? | None | 1 to 4 a week | 5+ a week |
| **Q40:** Are you vegan (completely avoid animal products including meat, fish, eggs and dairy)? | No | | Yes |

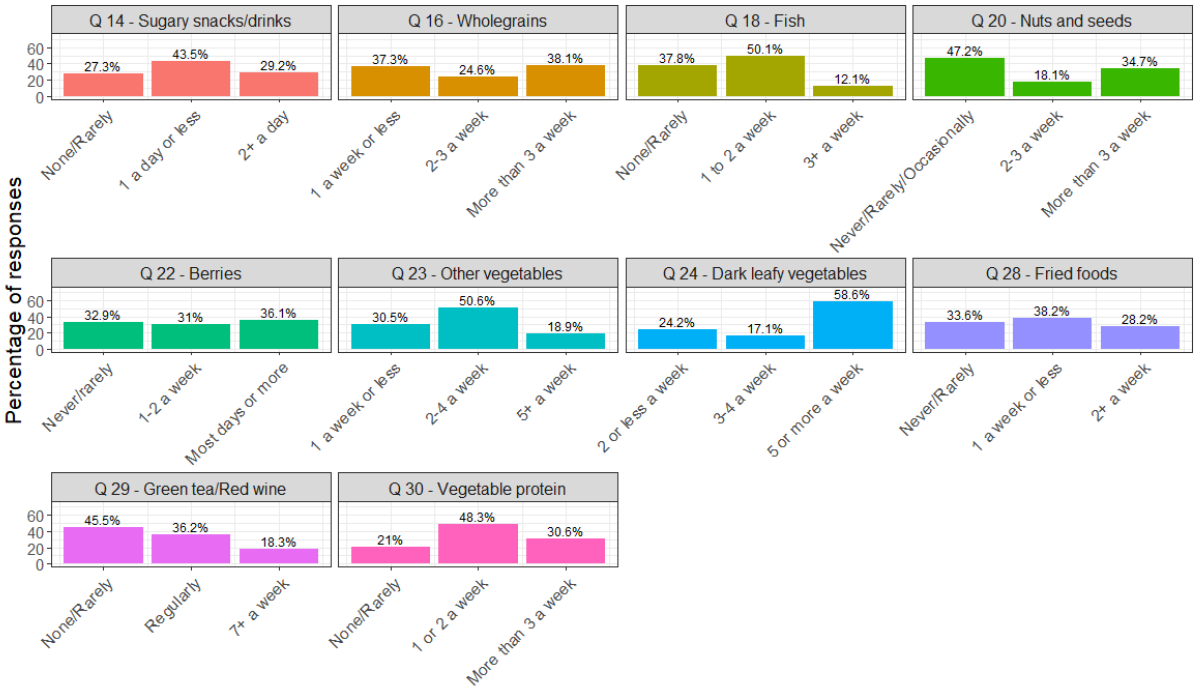


**S5. Supplementary Figure 3.** Distributions of participants' responses to the 10 queries selected for the calculation of individual MIND* scores.

### S6. Comparison between elastic net regression and linear regression models performance

A direct comparison between mean-squared errors (MSE) and R-squared statistics from ordinary least square regression (OLS) and elastic net regression (ENR) models pointed to similar performances compared to baseline, regardless of alpha level, as shown in Supplementary Table 3. The relative contribution of each nutrition-related or demographic question to participants' cognitive function under OLS and ENR models was also found to be very close. Such contributions, in absolute values, are presented in Supplementary Table 4. For simplicity, data are shown for a representative subset of alpha levels only (i.e. alpha = 0, 0.1, 0.5 and 1). A graphic representation of the relative contribution of the top 10 predictors identified by the ENR algorithm at alpha = 0.5 is shown in Supplementary Figure 4. Estimated contributions resulting from the OLS model for these same predictors are also shown to facilitate direct comparisons between model performances.

**Supplementary Table 3:** Mean-squared errors and R-squared statistics for baseline, OLS and ENR models at various alpha levels.

| | **MSE** | **$R^2$** |
|---|---|---|
| Baseline | 156.35 | 0.0004 |
| OLS | 132.16 | 0.1546 |
| ENR - 0 | 132.15 | 0.1548 |

| | | |
|---|---|---|
| ENR – 0.1 | 132.16 | 0.1547 |
| ENR – 0.2 | 132.16 | 0.1547 |
| ENR – 0.3 | 132.17 | 0.1547 |
| ENR – 0.4 | 132.17 | 0.1547 |
| ENR – 0.5 | 132.17 | 0.1547 |
| ENR – 0.6 | 132.18 | 0.1546 |
| ENR – 0.7 | 132.17 | 0.1547 |
| ENR – 0.8 | 132.17 | 0.1547 |
| ENR – 0.9 | 132.17 | 0.1547 |
| ENR - 1 | 132.17 | 0.1547 |

**Supplementary Table 4**: Relative absolute contribution of model predictors to cognitive function for OLS and selected ENR models (alpha = 0, 0.1, 0.5, 1).

| | **OLS** | **ENR - 0** | **ENR – 0.1** | **ENR – 0.5** | **ENR - 1** |
|---|---|---|---|---|---|
| Age | 3.0311 | 2.9032 | 3.0174 | 3.0276 | 3.0290 |
| Education | 0.1492 | 0.1446 | 0.1482 | 0.1486 | 0.1487 |
| Gender | 1.0246 | 1.0222 | 1.0195 | 1.0201 | 1.0205 |
| Q14 | 0.6232 | 0.5301 | 0.5409 | 0.5532 | 0.5592 |
| Q15/32 | 0.7664 | 0.5761 | 0.6360 | 0.6585 | 0.6682 |
| Q16 | 2.3458 | 2.1581 | 2.2891 | 2.3079 | 2.3129 |
| Q17 | 6.19508 | 5.8478 | 6.1360 | 6.1659 | 6.1714 |
| Q18/19 | 1.96678 | 1.8740 | 1.9183 | 1.9291 | 1.9331 |
| Q20 | 0.8573 | 0.7956 | 0.8141 | 0.8221 | 0.8257 |
| Q21 | 0.7612 | 0.6908 | 0.7066 | 0.7180 | 0.7231 |
| Q22 | 0.5696 | 0.5350 | 0.5122 | 0.5191 | 0.5236 |
| Q23 | 1.2995 | 1.2626 | 1.2642 | 1.2697 | 1.2727 |
| Q24/27 | 1.8744 | 1.7507 | 1.8319 | 1.8452 | 1.8492 |
| Q28 | 1.7038 | 1.6652 | 1.6600 | 1.6663 | 1.6700 |
| Q29 | 2.0245 | 1.9186 | 1.9890 | 1.9997 | 2.0027 |
| Q30 | 1.3100 | 1.2542 | 1.2626 | 1.2697 | 1.2733 |
| Q31 | 4.5297 | 3.9988 | 4.3911 | 4.4410 | 4.4524 |
| Q34 | 1.0437 | 1.0193 | 1.0313 | 1.0341 | 1.0352 |
| Q38 | 2.0445 | 1.9618 | 2.0114 | 2.0202 | 2.0231 |
| Q39 | 0.4787 | 0.3296 | 0.3610 | 0.3797 | 0.3883 |
| Q40 | 0.5009 | 0.4392 | 0.4872 | 0.4935 | 0.4949 |

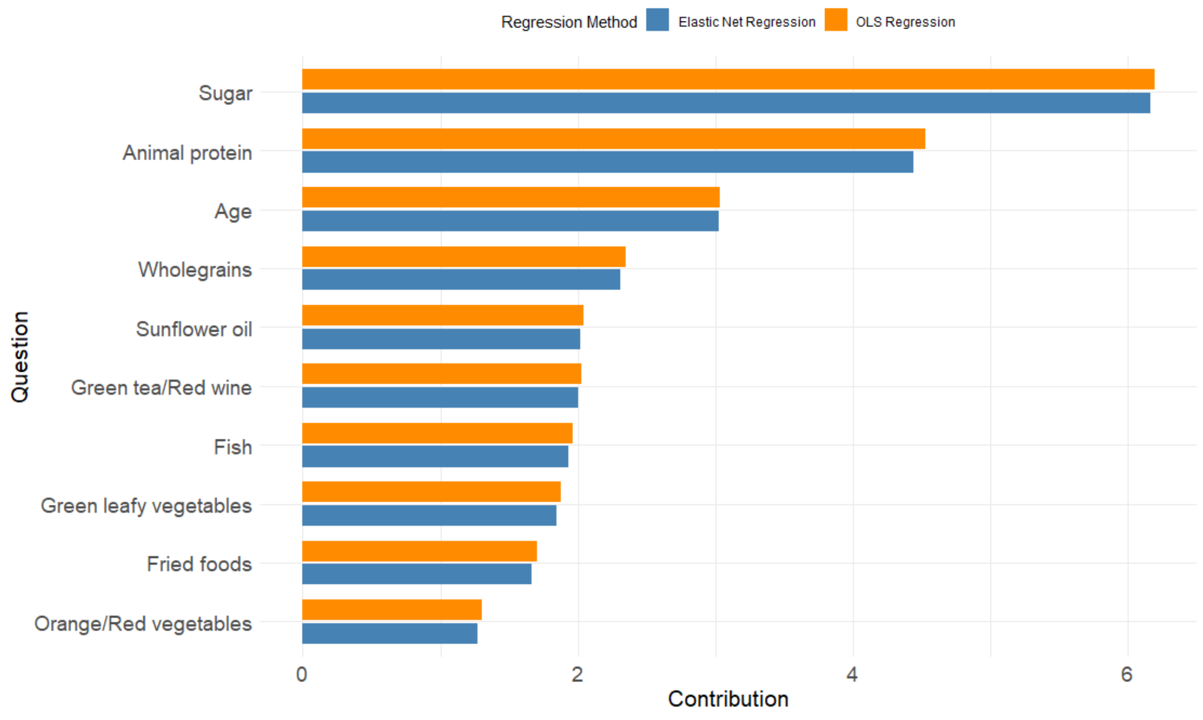


**Supplementary Figure 4.** Top 10 predictors of cognitive function identified by the ENR (alpha = 0.5) and OLS regression models.

*Note:* Displayed predictors correspond to the top 10 variables identified by the ENR algorithm at alpha = 0.5 and are presented in descending order of relative contribution to cognitive function.

### S7. Supplementary Table 5: Survey questions and linear scales used to calculate the MIND* Linear score following the original MIND questions.

| Question | 0 | 0.5 | 1 |
|---|---|---|---|
| **Q14:** How many sugar-based snacks or drinks (biscuits, cakes, chocolate, sweets, fizzy drinks, fruit juices) do you consume each day? | 2+ a day | 1 a day or less | None/Rarely |
| **Q16**: How many times a week do you eat wholegrains? | 1 a week or less | 2-3 a week | 3+ a week |
| **Q18:** How many times a week do you eat fish? | Never/Rarely | 1-2 a week | 3+ a week |
| **Q20:** How frequently do you eat raw walnuts, pecans, macadamia, chia or flax seeds? | Never/Rarely/Occasionally | 2-3 a week | 3+ a week |
| **Q22**: How many servings of berries, cherries, plums, pomegranate or their juice do you have a day? | None/Rarely | 1-2 a week | Most days or more |
| **Q23**: How many servings of orange or red vegetables do you have a week? | 1 a week or less | 2-4 a week | 5+ a week |
| **Q24**: How many servings of green leafy or cruciferous vegetables do you have a week? | 2 a week or less | 2-4 a week | 5+ a week |
| **Q28:** How many times a week do you eat fried, deep fried or browned foods including crisps, chips and fried take away food? | 2+ a week | 1 a week or less | Never/Rarely |

| | | | |
|---|---|---|---|
| **Q29:** How many green or herbal teas or small glasses (175mls) of red wine do you drink a week? | None/Rarely or 7+ a week | Regularly | |
| **Q30:** How many times a day do you eat vegetable protein (e.g. beans, lentils, tofu, quinoa, peas, corn)? | Never/Rarely | 1-2 a week | 3+ a week |

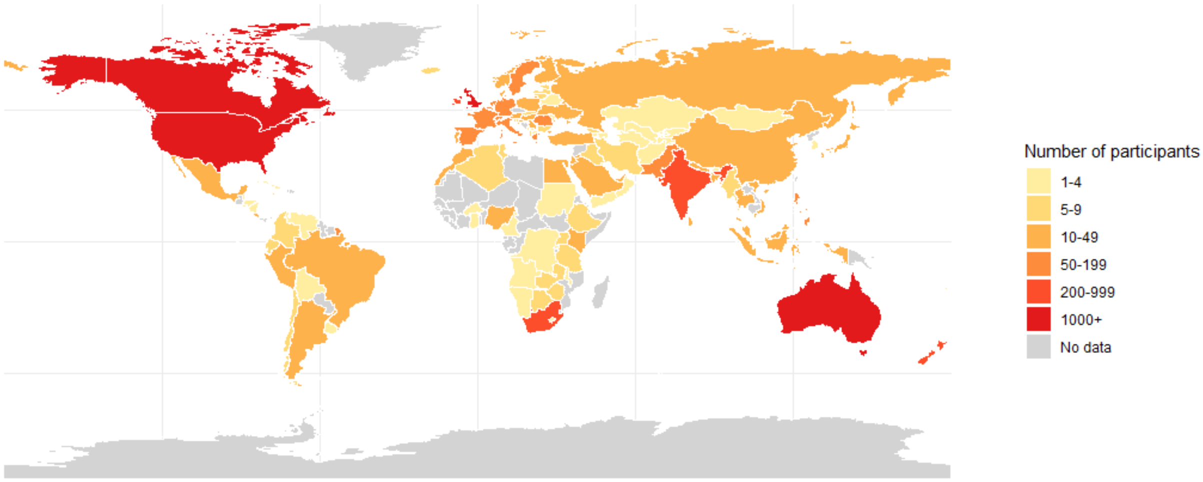


**S8. Supplementary Figure 5**: World map of study participants providing cultural background information.

The majority of participants for this study who provided cultural background information was from the United Kingdom (47.93%), followed by the United States of America (21.35%), Australia (6.54%) and Canada (6.35%). A further 135 countries cumulatively contributed the remaining 17.83% of responses, with 79 of these countries including fewer than 10 participants each.

### S9. Distribution of responses to questions 18, 22 and 28 by education group.

In the attempt to understand whether socio-economic status may have influenced the counterintuitive results we observed in relation to weekly consumption of fish, berries and fried foods, we looked at the distribution of participants' responses based on the number of years of full-time education they completed. The cutoff points we set for this qualitative analysis were 12 and 17 years, corresponding to the first and third quartile of data for this demographic variable, respectively.

As shown in Supplementary Figures 6, 7 and 8, respondents sitting at the lower end of the education spectrum tended to consume fish and berries more sporadically than their more educated counterparts, whereas the opposite was true in regard to fried foods. While it is important to acknowledge that the number of years of full-time education does not necessarily translate to higher or lower incomes in a linear fashion, these observations suggest that participants' socio-economic status might have played a role in driving our unexpected results and therefore warrant further investigations incorporating more granular data on respondent's financial well-being.

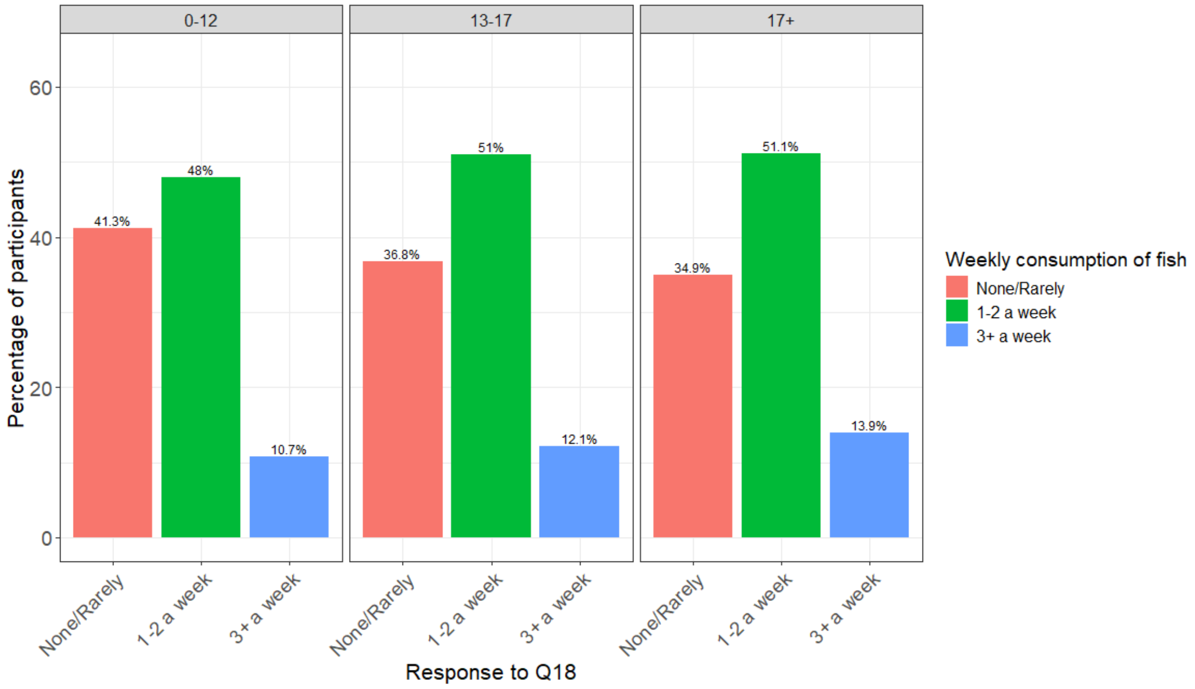


**Supplementary Figure 6.** Distribution of participants' responses to question 18 (fish) by education group.

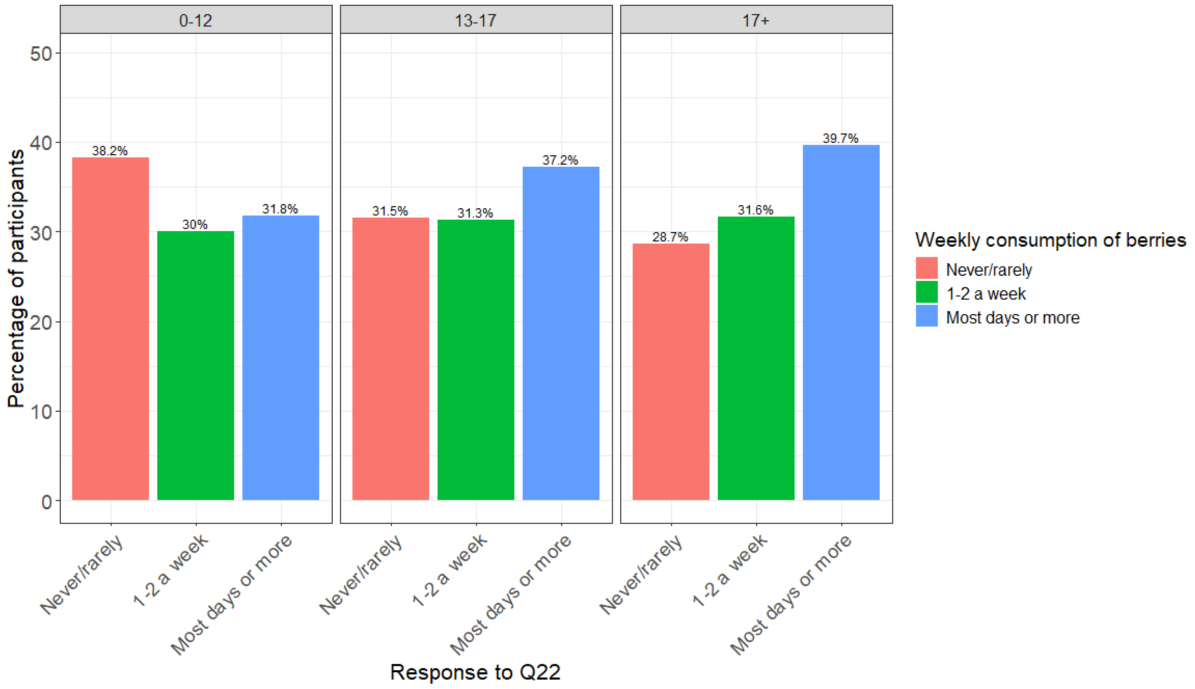

**Supplementary Figure 7.** Distribution of participants' responses to question 22 (berries) by education group.

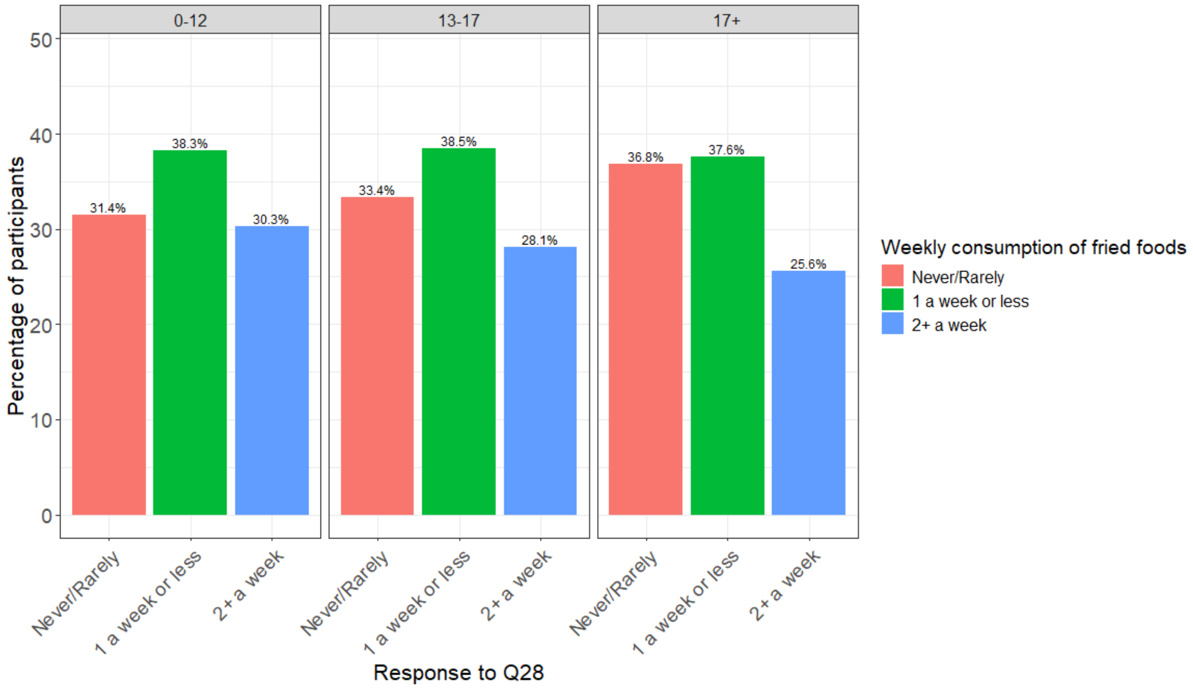


**Supplementary Figure 8.** Distribution of participants' responses to question 28 (fried foods) by education group.

### S10. ENR regression coefficients including interaction terms with age.

We conducted an additional ENR analysis by including interaction terms between age and nutrition-related variables. The main predictors of cognitive function identified by the algorithm remained largely the same as in previous analyses (see Supplementary Figure 9). In fact, across the whole spectrum of our selected variables, main effects were found to be much larger compared to interaction terms. To highlight these findings and facilitate their interpretation, the absolute regression coefficients of the dummy variables corresponding to each nutrition-related question have been summed, both for main effects and interaction effects. These cumulative contributions are presented side-by-side in Supplementary Table 6.

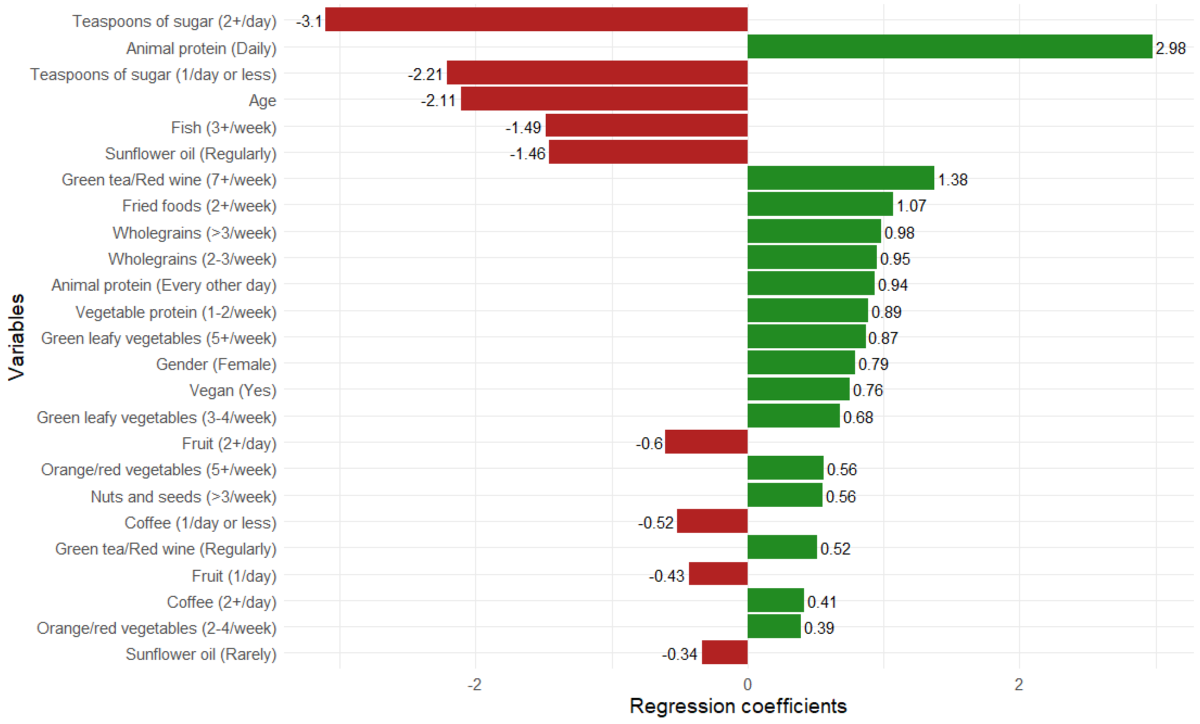


**Supplementary Figure 9.** Top 25 predictors of cognitive function identified by the ENR (alpha = 0.5) including interaction terms with age.

**Supplementary Table 6.** Cumulative sums of absolute regression coefficients for main and interaction effects obtained from the mixed effects ENR.

| Variable | Main effect coefficient | Interaction coefficient |
|---|---|---|
| Q14 [Sugary snacks/drinks] | 0.3589 | 0.0109 |
| Q15/32 [Refined carbohydrates] | 0.1413 | 0.0028 |
| Q16 [Wholegrains] | 1.9335 | 0.0509 |
| Q17 [Teaspoons of sugar] | 5.3145 | 0.1441 |
| Q18/19 [Fish] | 1.7429 | 0.0285 |
| Q20 [Nuts and seeds] | 0.7486 | 0.0142 |
| Q21 [Fruit] | 1.0354 | 0.0456 |
| Q22 [Berries] | 0.3877 | 0.0705 |
| Q23 [Orange/Red vegetables] | 0.9496 | 0.0355 |
| Q24 [Green leafy vegetables] | 1.5486 | 0.0043 |
| Q28 [Fried foods] | 1.2617 | 0.0638 |
| Q29 [Green tea/Red wine] | 1.8923 | 0.1139 |
| Q30 [Vegetable protein] | 1.1249 | 0.0041 |
| Q31 [Animal protein] | 3.9170 | 0.0305 |
| Q34 [Coffee] | 0.9326 | 0.1162 |
| Q38 [Sunflower oil] | 1.7940 | 0.0673 |
| Q39 [Eggs] | 0.4164 | 0.0000 |
| Q40 [Vegan diet] | 0.7553 | 0.0725 |
